# Integrative Genomic and Immune Repertoire Profiling Identifies Clonal Signatures Linked to Antithyroid Drug-Induced Agranulocytosis

**DOI:** 10.1101/2025.11.27.25341185

**Authors:** Yi-Hui Huang, Sheng-Kai Lai, Yu-Hsuan Yang, Jun-Liang Chang, I-Hsuan Chiu, Szu-Chieh Chen, Po-Sheng Lee, Chia-Hung Lin, Wei-Yih Chiu, Wan-Chen Wu, Jin-Ying Lu, Chih-Yuan Wang, Jacob Shujui Hsu, Chien-Yu Chen, Chia-Lang Hsu, Ya-Chien Yang, Wei-Shiung Yang, Chun-Jui Huang, Shyang-Rong Shih, Pei-Lung Chen

**Author notes:** Correspondence: Chun-Jui Huang, Shyang-Rong Shih, Pei-Lung Chen, #288667. These authors contributed equally to this work (Yi-Hui Huang, Sheng-Kai Lai).

## Abstract

Graves’ disease (GD) is the leading cause of hyperthyroidism and is often treated with antithyroid drugs (ATDs). Although ATD therapy is effective, it might cause a rare but serious adverse effect called ATD-induced agranulocytosis (TiA), which can lead to severe neutropenia and life-threatening infections. Previous studies have shown that certain human leukocyte antigen (HLA) alleles, including *HLA-B*38:02* and *HLA-DRB1*08:03* in Asian populations, have been associated with TiA susceptibility. However, the underlying mechanisms remain unclear, highlighting the need to investigate the TiA-related immune alterations to better understand its pathogenesis and mechanisms.

In this study, we investigated the immune receptor repertoire in TiA patients. Global repertoire diversity, VJ gene usage, and V-J pairing remained preserved across phenotypes and disease phases. Notably, TiA patients exhibited several upregulated complementarity-determining regions 3 (CDR3) clonotypes compared to GD patients, suggesting their role in disease progression and pathogenesis. Single-cell immune repertoire analysis revealed that TiA-associated risk CDR3 sequences were predominantly expressed on CD8+ effector memory T cells (CD8 TEM) in patients with *HLA-B*38:02*, while CD4+ central memory T cells (TCM) showed increased expression of risk CDR3 sequences in patients with *HLA-DRB1*08:03*, suggesting distinct cellular mechanisms underlying HLA-associated TiA pathogenesis. In conclusion, this study sheds light on the adaptive immunoprofile associated with TiA development and provides insights into the adaptive immune profile of TiA and HLA-mediated disease susceptibility.

## INTRODUCTION

Graves’ disease (GD) is the leading cause of hyperthyroidism resulting from the production of autoantibodies that stimulate the thyroid gland. The global incidence of Graves’ disease is around 0.02-0.05% per year, with a higher prevalence in females^1^. The condition is often treated with antithyroid drugs (ATDs, also known as thionamide), such as propylthiouracil (PTU) and methimazole (MMI). However, a rare but serious adverse effect related to thionamide therapy is antithyroid drug-induced agranulocytosis (TiA) (defined as an absolute granulocyte count <500 mm^3^ within the first three months of ATD treatment), which can occur in 0.2–0.5% of GD patients receiving these medications^2^. Given that TiA is a severe condition that can lead to life-threatening infections, with an estimated mortality rate of 21%^3^, there is an urgent need to elucidate the pathogenic mechanisms by which ATD induce agranulocytosis and to identify potential biomarkers for detection and risk prediction.

Although the underlying mechanisms of TiA remain unclear, previous studies have indicated that TiA might be caused by two pathways: direct cellular toxicity and immune-mediated destruction. The direct mechanism involves oxidative damage when ATDs accumulate in neutrophils and are metabolized by myeloperoxidase or cytochrome P450 into reactive metabolites that trigger inflammasome activation^4^. The immune-mediated pathway involves circulating autoantibodies, including complement-dependent IgM antibodies, targeting mature granulocytes and myeloid progenitor cells, leading to complement-mediated opsonization and apoptosis^5, 6^. Importantly, bone marrow examination of 33 TiA patients has revealed that granulocytopoiesis is significantly impaired in TiA patients^7^. These findings suggest that TiA involves not only peripheral neutrophil destruction but also central suppression of granulocytopoiesis at the bone marrow level, indicating a complex pathogenic mechanism that combines both autoimmune and direct myelosuppressive effects.

Several drug-induced adverse effects have shown human leukocyte antigen (HLA) genetic predisposition, including carbamazepine (CBZ)-induced severe cutaneous adverse reactions (SCAR) (*HLA-B*15:02* and *HLA-A*31:01*)^8, 9^, allopurinol-induced SCAR (*HLA-B*58:01*)^10^, and abacavir-induced hypersensitivity syndrome (*HLA-B*57:01*)^11, 12^. Similarly, TiA has also been found to be associated with several HLA alleles (e.g., *HLA-B*38:02* and *HLA-DRB1*08:03* in Asian populations^13^, *HLA-B*27:05* in European populations^14^). Three complementary models have been proposed to explain HLA-linked drug adverse reactions: the hapten hypothesis, where drugs covalently bind to proteins forming immunogenic adducts presented by HLA molecules^15^; the pharmacologic interaction (p-i) model, proposing direct drug-HLA or drug-TCR binding without metabolic activation^16^; and the altered-repertoire model, where drugs alter HLA peptide-binding specificity, presenting novel self-peptides that might trigger autoimmunity^17^. Importantly, recent immune repertoire study of *HLA-B*15:02*-associated CBZ-induced SCAR has identified oligoclonal disease-causing TCR clonotypes, supporting the p-i model^18^. Moreover, given the importance of HLA in shaping immune receptors, individuals with different HLA alleles might exhibit distinct immunoprofiles and downstream immune responses^19, 20^. Therefore, profiling the adaptive immune receptor repertoire (AIRR) composed of T cell receptor (TCR) and B cell receptor (BCR), using high-throughput sequencing could provide novel insights into the pathogenesis and underlying mechanisms of HLA-related diseases.

In this study, we performed comprehensive immune repertoire profiling in Graves’ disease patients and those who developed ATD-induced agranulocytosis to elucidate the pathogenesis and underlying mechanisms of TiA. Previous studies have reported an increased risk of TiA in patients carrying *HLA-B*38:02* and *HLA-DRB1*08:03*, with estimated odds ratios (carriers to non-carriers) 21.48 and 6.13, respectively^13^. Moreover, the combined presence of *HLA-B*38:02* and *HLA-DRB1*08:03* leads to an elevated odds ratio of 48.41^13^. Based on these findings, patients enrolled in this study were assigned into four groups: (I) patients with only *HLA-B*38:02*, (II) those possessing only *HLA-DRB1*08:03*, (III) patients harboring both risk alleles, (IV) those lacking both alleles. Each group included patients with TiA and GD, as well as Controls. Additionally, we collected longitudinal samples from the same patients at different stages, including the acute phase (at diagnosis) and the remission phase (0.5-1 year post-diagnosis). By comparing the global repertoire composition and clonotypes across different disease phenotypes (TiA, GD, and Control) and disease phases (acute and remission phases), we aimed to uncover TiA-associated alterations in immune responses. Furthermore, integration of single-cell immune repertoire sequencing with whole transcriptomic data provided insights into cell type distribution and TCR sequences. Overall, our immune repertoire study identified TiA-associated risk clonotypes, shedding light on the mechanisms underlying TiA onset, pathogenesis, and disease progression.

## RESULTS

### HLA Genotyping Validation Confirms Previously Identified Susceptibility Alleles in TiA

HLA genotyping of 65 TiA patients, 167 GD patients, and 125 controls validated significant associations at previously identified *HLA-B*38:02* (*p* = 3.49 × 10⁻¹¹) and *DRB1*08:03* (*p* = 2.00 × 10⁻⁴) in TiA versus GD cases from our cohorts (Fig. 1B)^13, 21^. Subjects were stratified into four groups according to their HLA risk allele status for subsequent immune repertoire analysis: Group I carrying only *HLA-B*38:02* (B*38:02 only), Group II carrying only *HLA-DRB1*08:03* (DRB1*08:03 only), Group III carrying both risk alleles (Both), and Group IV lacking both alleles as controls (Without) (Table S1). Each group included TiA, GD, and Control samples that were analyzed by bulk TCR and BCR repertoire sequencing, with a subset additionally examined by single-cell TCR sequencing coupled with transcriptomic profiling for comprehensive characterization (Fig. 1C).

**Figure 1.**
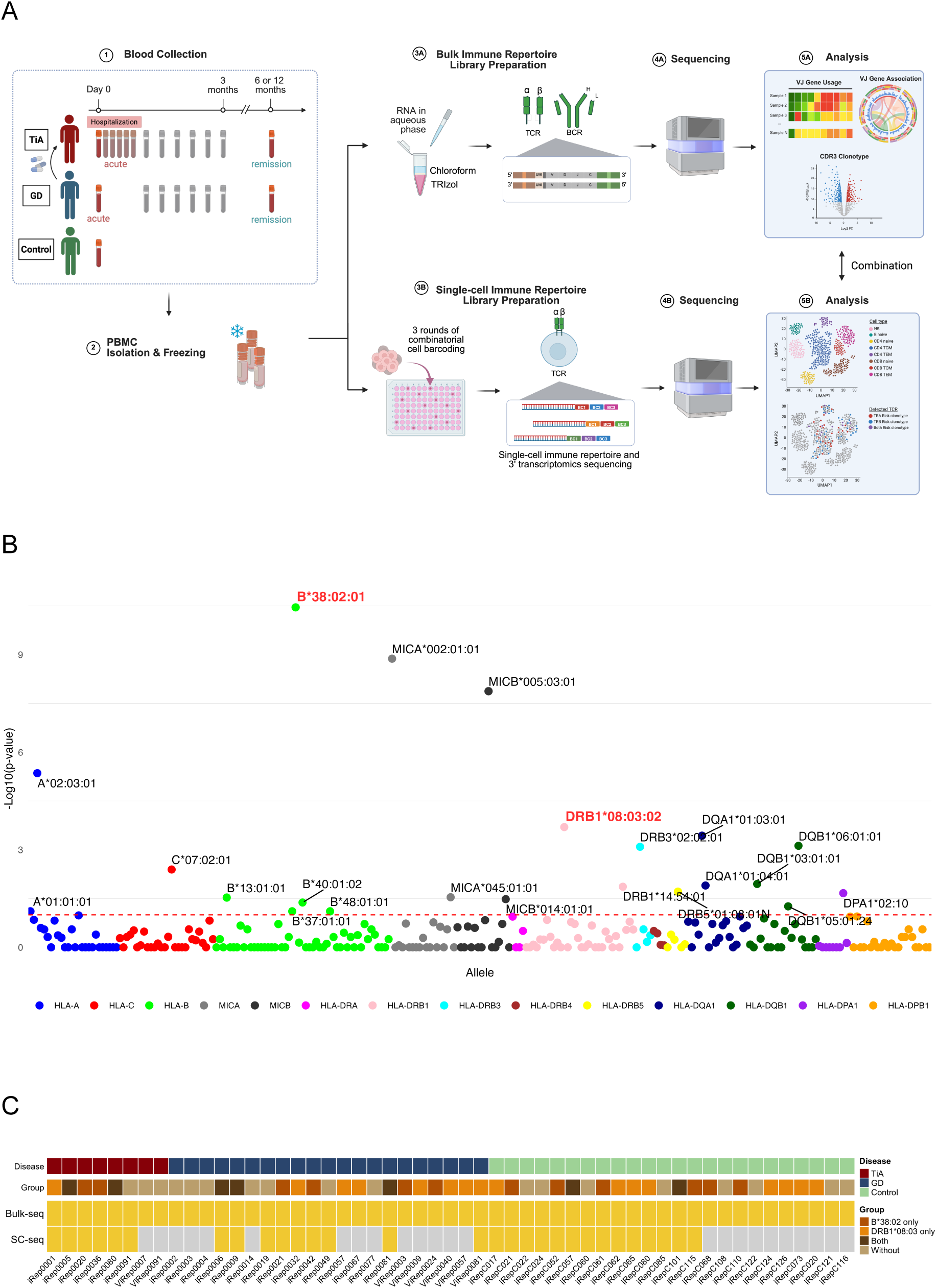
Experimental design overview and HLA allele association validation. (**A**) (1) Our study enrolled three cohorts: GD patients who developed TiA following ATD treatment (TiA), GD patients without TiA (GD), and normal controls (Control). For TiA and GD patients, longitudinal samples were collected at two time points: the acute phase (at initial diagnosis) and the remission phase (typically 0.5-1 year post-diagnosis). (2) Peripheral blood mononuclear cells (PBMCs) from all participants were isolated and subjected to both bulk and single-cell immune repertoire sequencing. (3A) For bulk sequencing, total RNA was extracted using TRIzol and chloroform, followed by cDNA synthesis, template switching, and PCR amplification to generate sequencing libraries. (3B) For single-cell sequencing, sequencing libraries were prepared by three rounds of combinatorial cell barcoding. (4-5) Following sequencing, both datasets were integrated for comprehensive comparative analyses. Workflow was created in BioRender (https://BioRender.com). (**B**) Manhattan plot depicting the distribution of HLA allele frequencies compared between TiA and GD patients in our cohort. Two previously reported TiA-susceptibility alleles (*B*38:02* and *DRB1*08:03*) are labeled in red. (**C**) Study participants and sequencing coverage across the study cohort. The top panel indicates disease phenotype (TiA: red; GD: blue; Control: green). The second panel shows HLA genotype groups based on the presence of TiA-associated alleles (*B*38:02* only: dark orange; *DRB1*08:03* only: orange; both alleles: brown; without either allele: light brown). The bottom two panels illustrate the availability of bulk immune repertoire sequencing (Bulk-seq) and single-cell immune repertoire sequencing (SC-seq) data for each sample (yellow: data available; gray: data unavailable).

### Repertoire-level Profiling Reveals Comparable Broad TCR and BCR Composition Across Disease States and Controls

We initially assessed three diversity indices of TCR and BCR repertoires to evaluate changes in diversity richness (relative D50), evenness (Shannon entropy), and clonal expansion (Simpson’s dominance). The distribution of these diversity metrics for each subject was visualized using tree maps (Fig. S1A-B). Subsequently, we compared diversity metrics across five groups: TiA patients in the acute phase (TiA-acute), TiA patients in remission (TiA-remission), GD patients in acute phase (GD-acute), GD patients in remission (GD-remission), and normal controls (Control).

For TCR repertoire, no statistically significant differences in diversity were observed among the five groups (Fig. 2A). To examine more detailed temporal dynamics within individuals, we further compared paired longitudinal samples across distinct disease phases. Paired comparisons of TCR repertoire revealed no differences between disease phases in TiA patients (Fig. 2B). For BCR repertoire, although GD patients during remission showed a significant reduction in Shannon entropy compared to normal controls, the other two diversity indices (relative D50 and Simpson dominance) showed no significant differences (Fig. 2C). Since these three indices reflect different aspects of repertoire diversity, the comprehensive evaluation using all three indices suggests that overall BCR repertoire diversity is similar across groups. Paired longitudinal analysis of BCR repertoire similarly showed no significant differences between disease phases in TiA patients (Fig. 2D). Collectively, these findings indicate that repertoire diversity does not differ significantly across disease phenotypes or disease phases.

**Figure 2.**
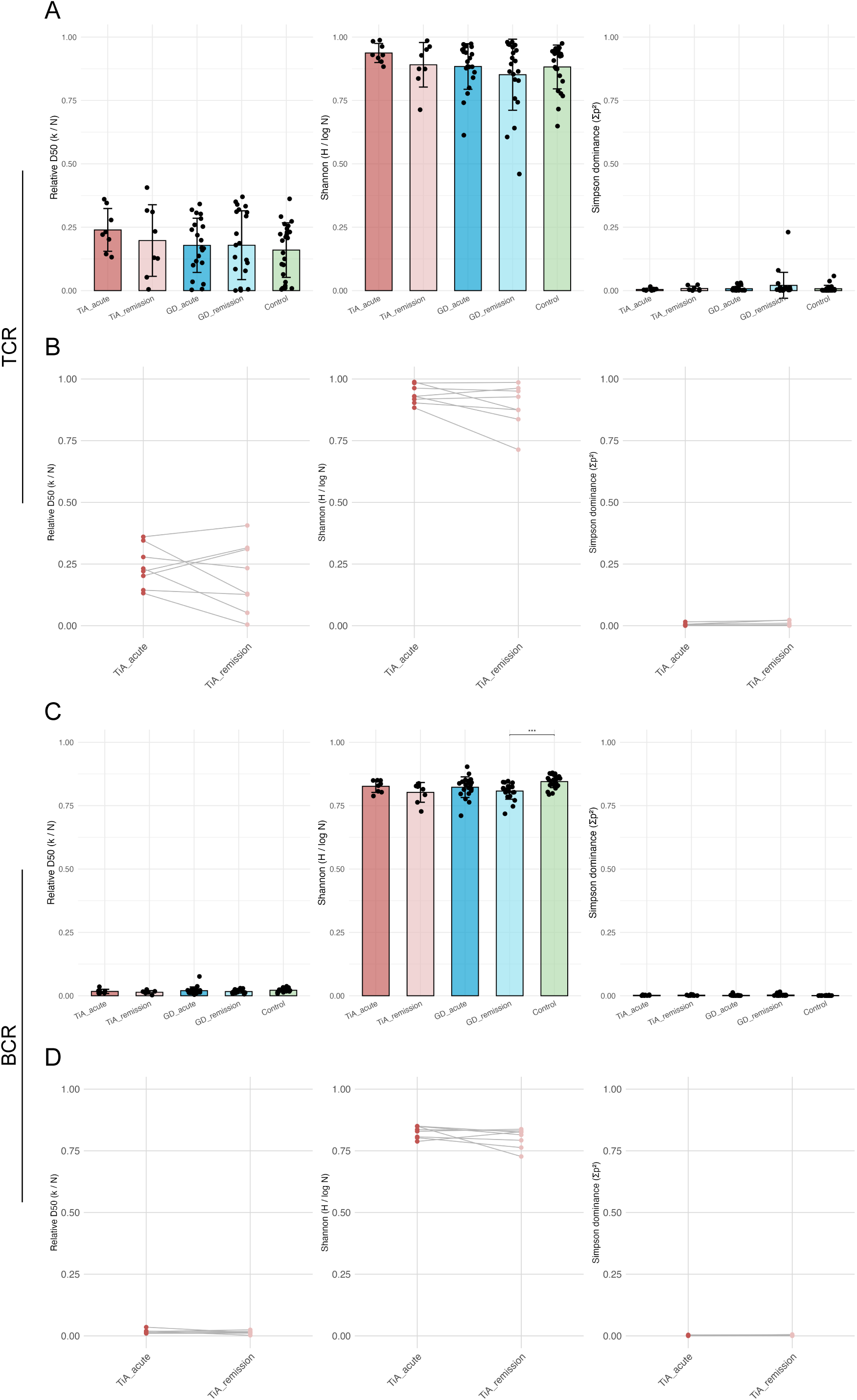
Immune repertoire diversity across disease phenotypes and phases. (**A**) Comparison of three diversity metrics (relative D50, Shannon entropy, and Simpson’s dominance index) of TCR repertoire across five groups. Each bar represents the mean value, and individual data points are overlaid. Statistical significance was assessed using pairwise two-sample t-tests with Benjamini–Hochberg (BH) correction for multiple comparisons. (**B**) Paired longitudinal comparisons of TCR repertoire diversity metrics between TiA-acute and TiA-remission patients. Gray lines connect paired samples, and statistical significance was determined by Wilcoxon signed-rank test and adjusted with BH correction for multiple testing. (**C**) Comparison of three diversity metrics of BCR repertoire across five groups, as described in (A). (**D**) Paired longitudinal comparisons of BCR repertoire diversity metrics between TiA-acute and TiA-remission patients, as described in (B). Statistical significance was determined as \**p* < 0.05, \*\**p* < 0.01, \*\*\**p* < 0.001.

V(D)J recombination generates immune repertoire diversity through the selection and joining of variable (V), diversity (D), and joining (J) gene segments. To further investigate general immune profile patterns across disease phenotypes and phases, we analyzed the V/J gene usage in five groups. Overall, V/J gene usage pattern of both TCR and BCR appeared similar between five groups (Fig. S2, left). While certain genes showed elevated frequencies in TiA-acute and TiA-remission patients, statistical comparison revealed no significant differences in the usages of these genes between groups (Fig. S2, right). Beyond gene selection preferences, V-J gene pairing also contribute to receptor diversity. We therefore analyzed V-J pairing patterns across the five groups. Comparison of V-J pairings revealed generally high overlap across the five groups. A notable exception was *TRAV-TRAJ*, which showed low similarity among most groups (Fig. S3A), while *TRBV-TRBJ*, *IGHV-IGHJ*, *IGKV-IGKJ*, and *IGLV-IGLJ* all maintained high cosine similarity (> 0.8) (Fig. S3B-E). This low *TRAV-TRAJ* similarity likely results from its larger combinatorial space (∼2,050) compared to *TRBV-TRBJ* (∼650)^22^, which promotes inter-individual variability. Consistently, the elevated *TRAV-TRAJ* frequencies in TiA were predominantly driven by individual samples (e.g., *TRAV5-TRAJ43* in iRep0036-1), suggesting that observed differences in *TRAV-TRAJ* pairing between groups are more likely driven by individual variation than disease-associated patterns (Table S2).

In summary, repertoire-level analyses revealed no significant differences in diversity, V/J gene usage, or overall V-J pairing patterns between disease phenotypes and phases, suggesting that TCR and BCR repertoire compositions might not be altered by TiA onset and progression.

### Validation of Clonotype-level Approach Through GD-associated Risk CDR3 Similarity to Pathogenic Antibodies

Despite analyzing repertoire-level characteristics including diversity, V/J gene usage, and V-J pairing patterns across the five groups, we observed no significant differences between groups, particularly between TiA-acute and GD-acute patients. This suggests that TiA-associated immune responses may involve subtle, clonotype-specific alterations rather than broad repertoire-wide shifts. We therefore performed clonotype-level analysis focusing on CDR3 regions, which directly determine antigen specificity. Following the concept of ’public T/BCRs’—shared immune receptor sequences that provide disease insights^18, 23^—we aimed to identify TiA-associated CDR3 clonotypes (referred to as clonotypes) that may be involved in TiA pathogenesis.

Given that TiA-associated antigens remain poorly understood, we validated our analytical pipeline by first identifying GD-associated clonotypes and assessing their predicted binding affinity to the known GD-associated antigen, thyroid-stimulating hormone receptor (TSHR)^24^.

We examined IGK clonotypes in GD-acute patients and normal controls using two parallel approaches. First, frequency-based analysis identified clonotypes enriched (Log_2_ fold-changes (FC) > 0, *p* < 0.05, Wilcoxon rank-sum test) and decreased (Log2FC < 0, *p* < 0.05) in GD-acute patients (Fig. S4A). Second, incidence-based analysis applied Fisher’s exact test to identify clonotypes with significant incidence differences (*p* < 0.05), which were further classified as GD-specific (incidence = 0 in all controls) or control-specific (incidence = 0 in all GD-acute patients) (Fig. S4B). The overlap between frequency- and incidence-based candidates was examined using Venn diagrams (Fig. S4C-D). Notably, all incidence-based GD-acute-specific clonotypes (n=140) were also identified by frequency-based analysis, suggesting that clonotypes exclusively present in GD-acute patients also tend to be enriched in frequency (Fig. S4C). To prioritize candidates for structural analysis, clonotypes identified by both approaches were further filtered by a stricter incidence threshold (*p* < 0.01) and selected for subsequent antibody-antigen docking.

We further performed structural modeling and docking analysis to characterize the binding affinity of GD-associated clonotypes. GD-associated risk clonotypes (Fig. S4E) and protective clonotypes (Fig. S4F) were first clustered based on CDR3 sequence similarity using BLOSUM62-based hierarchical clustering (distance cutoff = 16), a widely adopted approach in immune repertoire studies^25–27^, as sequence similarity reflects similar loop conformations and potential shared epitope recognition patterns. The structures of risk and protective clonotypes (IGH derived from the reference structure) were then generated and docked with TSHR using ClusPro^28–32^. For each docking model, we calculated epitope coverage relative to the reference structure and binding energy using PRODIGY^33, 34^. Docking results revealed that risk clonotypes demonstrated significantly higher epitope coverage compared to protective clonotypes (Fig. S4G), indicating greater similarity to the reference binding site. Although binding energies showed no significant difference between risk and protective clonotypes (Fig. S4H), the risk clonotype “CQQRSSWPQTF” with optimal combined scores (high epitope coverage and low binding energy) displayed a binding pose highly similar to the reference autoantibody (Fig. S4I). In contrast, the protective clonotype “CQQSNTVPYTF” showed a distinct binding pose from the reference (Fig. S4J). Overall, docking analysis revealed that risk clonotypes identified from our repertoire analysis exhibited structural and binding characteristics consistent with pathogenic thyroid-stimulating autoantibodies.

### Clonotype-level Analysis Identifies HLA-linked TiA-associated TCR CDR3 Clonotypes

After validating our clonotype-level analytical pipeline with GD-associated clonotypes, we applied it to identify TiA-associated risk clonotypes that are especially present in the acute phase. Previous studies have shown that diseases with genetic predisposition to HLA class I alleles are often associated with CD8+ T cell responses^18^, while those associated with HLA class II alleles typically involve CD4+ T cells and B cells^35^. Since TiA risk HLA alleles include both HLA class I and class II^13^, we examined risk clonotypes in both the TCR and BCR repertoire.

To identify risk clonotypes specifically related to the acute phase in TiA, we compared clonotype frequencies and incidences of TCR (Fig. 3 and Fig. S5) and BCR (Fig. S6) between TiA-acute and GD-acute patients. For TiA-associated risk TRA and TRB clonotypes, frequency-based analysis identified enriched clonotypes (Fig. 3A-B), while incidence-based analysis identified TiA-acute-specific clonotypes (Fig. S5A-B). Notably, TiA-associated risk clonotypes identified by the incidence-based approach were also frequency-enriched (Fig. S5C-D), suggesting that these clonotypes undergo clonal expansion rather than merely appearing in TiA-acute patients. Similar patterns were observed in BCR repertoires, where incidence-based risk clonotypes were fully encompassed within frequency-enriched clonotypes across IGH, IGK, and IGL chains (Fig. S6).

**Figure 3.**
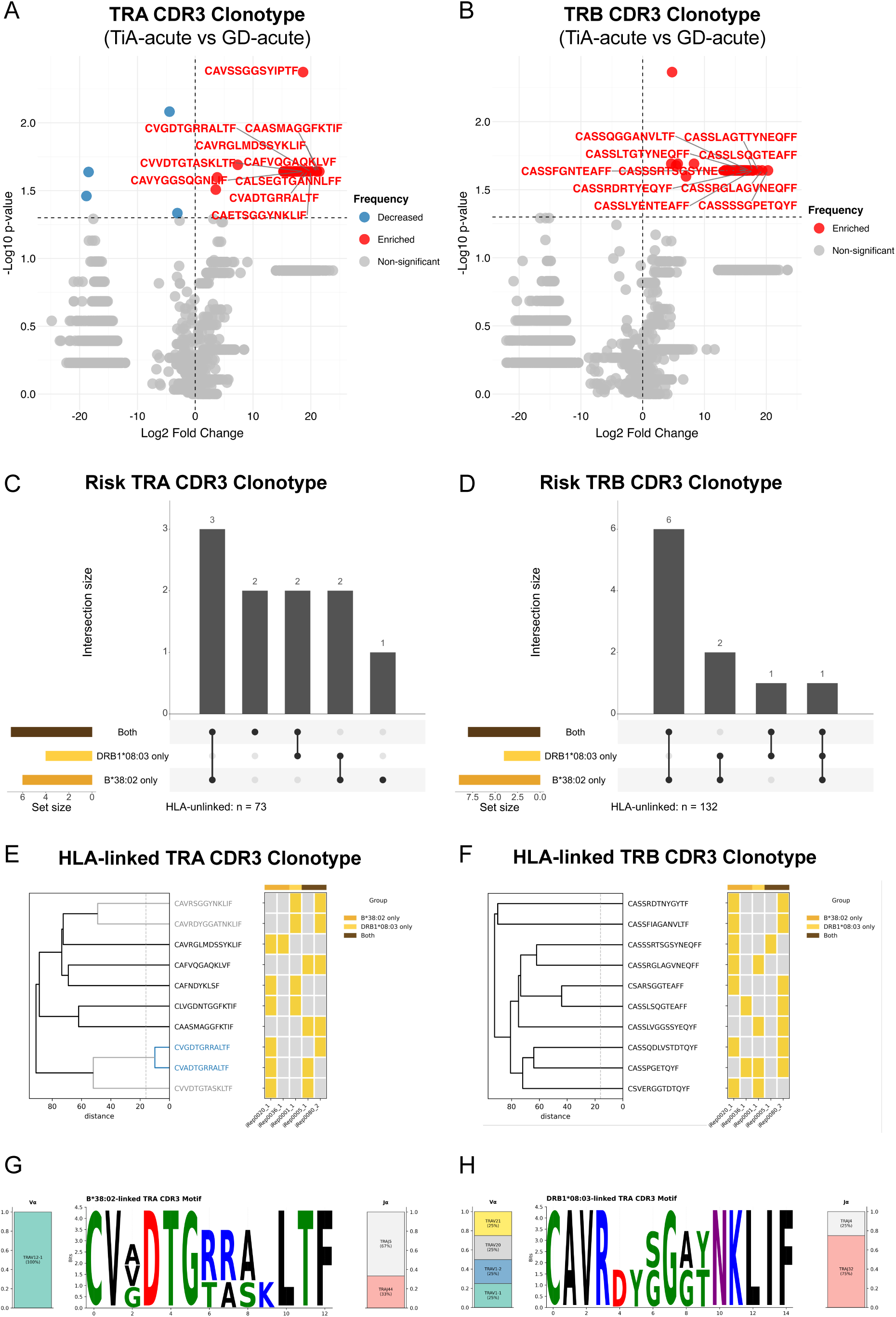
Identification of TiA-associated TCR risk clonotypes and HLA-linked CDR3 patterns. (**A-B**) Volcano plots showing CDR3 clonotypes of TRA (A) and TRB (B) with Log_2_ FC versus -Log_10_ (*p*-value) in TiA-acute patients compared to GD-acute subjects. Statistical significance was calculated using Wilcoxon rank-sum test. Red dots indicate significant risk clonotypes (*p* < 0.05 and Log_2_ FC > 0); blue dots indicate significant protective clonotypes (*p* < 0.05 and Log_2_ FC < 0); and gray dots represent non-significant clonotypes. CDR3 sequences of the most significant clonotypes are labeled. (**C-D**) The distribution of HLA-linked TRA (C) and TRB (D) CDR3 clonotypes in different HLA groups (*B*38:02* only, *DRB1*08:03* only, and Both). (**E-F**) Hierarchical clustering of HLA-linked TRA (E) and TRB (F) CDR3 clonotypes based on sequence similarity, displayed as dendrograms with distance on the x-axis. Each clonotype is annotated by its HLA group association (B*38:02 only, DRB1*08:03 only, or Both). Clonotypes forming a convergent cluster (distance < 16) are highlighted in blue, with their immediate sequence neighbor highlighted in gray. (**G-H**) Position weight matrices (PWM) of CDR3 sequences for TRA clonotypes linked to *HLA-B*38:02* (G) and *HLA-DRB1*08:03* (H). Each PWM depicts the V gene (left) and J gene (right) usage frequencies, with CDR3 amino acid sequences (middle). Amino acids are colored by chemical properties: acidic (red), basic (blue), hydrophobic (black), neutral (purple), and polar (green).

Although shared immune receptor sequences across individuals are expected to be exceedingly rare given the high diversity of the immune repertoire^36^, our identified TiA-associated risk clonotypes were present in at least two TiA-acute patients with increased average frequencies, rather than reflecting clonal expansion in single individuals (Table S3). These features are consistent with public clonotype patterns, further supporting their association with TiA pathogenesis.

Given the strong evidence for HLA allele predisposition in TiA^13^ and the established mechanism whereby HLA-related drug adverse reactions are mediated by interactions between public TCRs, drugs, and HLA risk alleles^17^, we sought to identify HLA-dependent patterns among TiA-associated TCR risk clonotypes identified in Fig. 3A-B. Since HLA-linked risk clonotypes are enriched during the acute phase^18^, we classified risk clonotypes according to the HLA group of TiA-acute patients carrying each clonotype: those present in the Without group were classified as HLA-unlinked, while others were defined as HLA-linked. Ten HLA-linked clonotypes were identified in both TRA and TRB, with 8/10 showing specificity to either B*38:02-carrying patients (B*38:02 only or Both) or DRB1*08:03-carrying patients (DRB1*08:03 only or Both) (Fig. 3C-D), suggesting HLA-dependent clonotype selection in TiA pathogenesis.

To further characterize these HLA-linked clonotypes, we examined CDR3 sequence similarity through hierarchical clustering (Fig. 3E–F). Within the TRA repertoire (Fig. 3E), CVGDTGRRALTF and CVADTGRRALTF formed a convergent cluster (distance < 16, highlighted in blue), with their neighboring clonotype CVVDTGTASKLTF, all three demonstrating *HLA-B*38:02*-favored pattern. In addition, CAVRSGGYNKLIF and CAVRDYGGATNKLIF, while not constituting a discrete convergent cluster, exhibited relatively closer sequence proximity compared to other clonotypes and both showed *HLA-DRB1*08:03* preference. Within the TRB repertoire, no convergent clusters were identified among the HLA-linked clonotypes (Fig. 3F). Position weight matrix analysis applied to TRA HLA-linked clonotypes revealed distinct sequence motifs for *HLA-B*38:02*-linked (Fig. 3G) and *HLA-DRB1*08:03*-linked (Fig. 3H) clonotypes; an equivalent analysis was not performed for TRB as no convergent clusters were identified among HLA-linked clonotypes.

To confirm that these HLA-linked clonotypes were not driven by germline-level bias, we verified that the V/J genes composing TiA-associated risk clonotypes were also carried by GD and control subjects at the germline level (Fig. S5E-F).

Overall, the identified HLA-specific clonotype signatures implicate HLA-TCR interactions as a mechanistic link between genetic predisposition and adaptive immune responses in TiA pathogenesis.

### Integration of Single-cell Immune Repertoire Uncovers HLA-specific Cellular Distribution of TiA-associated Risk TCR Clonotypes

To gain deeper insights into TiA-associated risk TCR clonotypes, we integrated single-cell TCR repertoire data with whole transcriptomic profiles. We clustered and visualized the overall cellular landscape using uniform manifold approximation and projection (UMAP) based on transcriptomic profiles. Initial annotation revealed the cell composition (Fig. 4A), and cells were subsequently colored according to the expressing status of TiA-associated risk clonotypes (Fig. 4B). Among cells expressing TiA-associated risk clonotypes, TCRs were detected mainly on CD8+ effector memory T cells (CD8 TEM, 36.4%), CD4+ central memory T cells (CD4 TCM, 29.2%), and naïve CD8+ T cells (CD8 Naïve, 24.0%) (Fig. 4C). Upon filtering for cells expressing TiA-associated risk clonotypes harbored by TiA-acute patients, risk clonotypes were predominantly expressed on CD8 TEM cells (57.7%), CD4 TCM (19.2%), CD8 Naïve (7.7%) (Fig. 4D). To further dissect the HLA-related patterns, we stratified these risk clonotypes according to patient HLA status. We defined B*38:02 carriers as patients carrying *HLA-B*38:02* (including B*38:02 only and Both groups), and DRB1*08:03 carriers as patients carrying *HLA-DRB1*08:03* (including DRB1*08:03 only and Both groups). Our analysis revealed distinct cellular preferences: risk clonotypes identified in B*38:02 carriers were expressed on CD8 TEM (62.5%) (Fig. 4E). In contrast, while risk clonotypes from DRB1*08:03 carriers remained predominantly expressed on CD8 TEM, there was a relatively higher proportion of CD4 TCM expression (15.4%) compared to the B*38:02 carriers (12.5%) (Fig. 4F).

**Figure 4.**
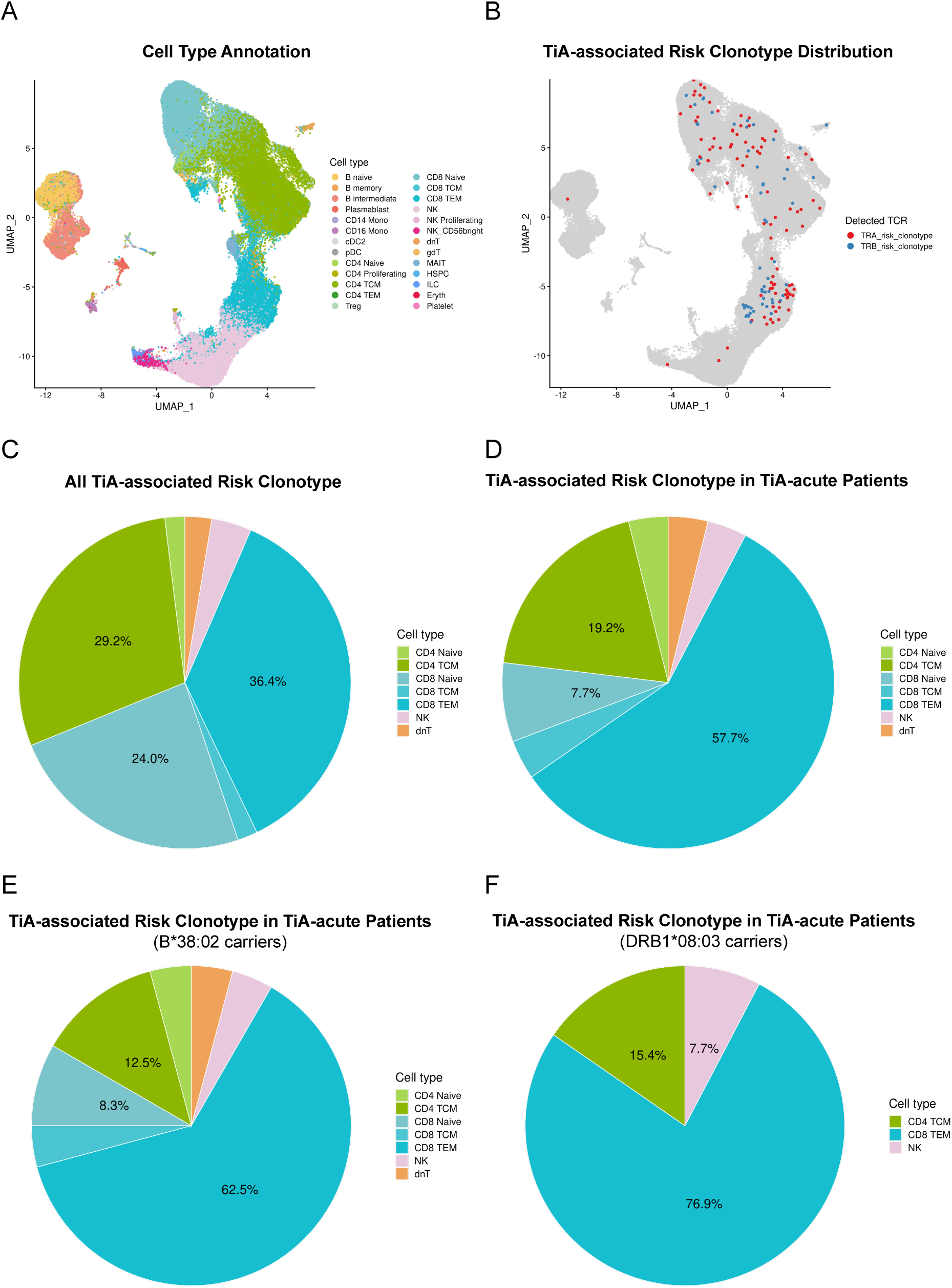
Single-cell immune profiling revealing the cellular distribution of TiA-associated risk CDR3 sequences. (**A-B**) UMAP visualization based on transcriptomic profiles was annotated according to cell type (A) and risk TCR clonotype detection (B). (**C-D**) Cell type composition among all cells expressing TiA-associated risk CDR3 clonotypes (C) and those harbored by TiA-acute patients (D). (**E-F**) Cell type composition of risk CDR3 clonotypes identified in patients carrying *HLA-B*38:02* (E) and *HLA-DRB1*08:03* (F). Cell types include: naïve B cell (B Naïve), intermediate B cell (B intermediate), memory B cell (B memory), plasmablast, CD14+ monocyte (CD14 Mono), CD16+ monocyte (CD16 Mono), conventional dendritic cell 2 (cDC2), plasmacytoid dendritic cell (pDC), naïve CD4+ T cell (CD4 Naïve), CD4+ proliferating T cell (CD4 Proliferating), CD4+ central memory T cell (CD4 TCM), CD4+ effector memory T cell (CD4 TEM), regulatory T cell (Treg), naïve CD8+ T cell (CD8 Naïve), CD8+ central memory T cell (CD8 TCM), CD8+ effector memory T cell (CD8 TEM), CD56-dim natural killer (NK), proliferating natural killer cell (NK Proliferating), CD56-bright natural killer cell (NK_CD56bright), double-negative T cell (dnT), gamma-delta T cell (gdT), mucosal associated invariant T cell (MAIT), hematopoietic stem and progenitor cell (HSPC), innate lymphoid cell (ILC), erythroid cell (Eryth), platelet (Platelet).

These HLA-specific cellular distribution patterns demonstrate that risk clonotypes associated with different HLA alleles are expressed on distinct T cell subsets. The predominance of CD8 TEM cells in B*38:02 carriers and the increased CD4 TCM representation in DRB1*08:03 carriers suggest that both HLA class I and class II pathways may contribute to TiA pathogenesis through divergent cellular mechanisms.

## DISCUSSION

In this study, immune repertoire profiling across five disease groups revealed no significant differences in global repertoire composition. However, several enriched TCR and BCR clonotypes were identified in TiA-acute patients compared to GD-acute patients, which we refer to as TiA-associated risk clonotypes. Further analysis revealed distinct CDR3 sequence motifs associated with different HLA risk alleles. Additionally, integration of single-cell sequencing data demonstrated that these risk clonotypes were predominantly expressed on HLA-dependent cell types.

The high diversity of immune repertoires, created by V(D)J recombination along with nucleotide addition and deletion, results in a theoretical diversity of 10^18^ in humans^37^. While this might suggest minimal overlap between individuals, accumulating evidence suggests that public T cell and B cell receptors are commonly observed among patient groups and can serve as predictive markers for disease^23^. This indicates that patients may share CDR3 sequences relevant to disease pathogenesis, even when present at low frequencies in peripheral blood. This concept is reflected in our observation: while no significant differences were observed at the repertoire-level (Fig. 2 and Figs. S1-3), several TiA-associated risk clonotypes were successfully identified in TiA-acute patients (Fig. 3). These findings suggest that TiA-associated risk clonotypes present at subtle enrichment are insufficient to alter overall repertoire composition, but are detectable at the clonotype level where they may play a role in disease pathogenesis. The identification of these public, disease-associated risk clonotypes strengthens the biological relevance of our findings and supports the hypothesis of convergent immune responses in TiA pathogenesis.

While the pathogenic mechanisms of TiA remain unclear, Graves’ disease represents a well-established autoimmune condition mediated by autoantibodies targeting the thyrotropin receptor (TSHR). To validate the reliability of our dual clonotype-level analytical approach, we compared the frequency and incidence of BCRs between GD-acute patients and normal controls to identify GD-associated risk clonotypes and protective clonotypes. Comparison of TSHR protein binding between GD-associated risk and protective clonotypes revealed significantly higher epitope coverage by risk clonotypes relative to protective clonotypes (Fig. S4G). This validation supports the robustness of our clonotype-level approach and its biological relevance to disease pathogenesis.

Our integration of bulk and single-cell sequencing data revealed that TiA-associated risk clonotypes exhibited HLA-dependent cellular distribution patterns, with risk clonotypes identified in B*38:02 carriers enriched mainly in CD8 TEM cells and those in DRB1*08:03 carriers enriched in CD4 TCM cells, suggesting that these HLA alleles may contribute to TiA pathogenesis through distinct pathways (Fig. 4). Notably, despite distinct predominant cell localization patterns, both HLA alleles showed increased proportion of CD8 TEM cells, indicating that downstream immune responses leading to agranulocytosis likely involve CD8 TEM participation. This observation was consistent with a previous study showing that CD8+ T cells could induce inflammatory responses in various tissues, including bone marrow, leading to inhibition of granulocytopoiesis^38^. Moreover, studies have shown that the proportions of CD4+ and CD8+ effector memory T cells are increased in both peripheral blood and bone marrow of patients with aplastic anemia^39^. Given that aplastic anemia and drug-induced agranulocytosis might share similar pathophysiological mechanisms involving immune-mediated suppression of hematopoiesis, the predominant localization of TiA-associated risk clonotypes on CD8 TEM cells suggests a biologically relevant role for these cells in disease pathogenesis.

While this study provides novel insights into HLA-associated immune signatures in TiA, several limitations highlight the need for further investigation. First, several technical and biological factors might limit comparative analyses of immune repertoire. The majority of clonotypes exhibit very low frequencies within diverse repertoires, and immune repertoires are largely individualized due to technical and biological variation, making statistical comparisons challenging^40, 41^. Additionally, as demonstrated in other drug-induced reactions, disease-relevant immune cells may be enriched at tissue sites rather than peripheral blood^18^, potentially limiting our ability to capture the complete immune landscape. Furthermore, drug-induced immune responses might show less pronounced clonal expansion compared to acute infections, consistent with our repertoire-level observations. Similar to autoimmune conditions where pathogenesis involves complex immune dysregulation, patients with systemic lupus erythematosus (SLE) exhibit diverse but less clonally expanded B-cell receptor repertoires^42^. As TiA likely shares similar pathophysiological features, the subtle clonal alterations might be inherently difficult to detect through repertoire-level comparisons. Second, while our integration of single-cell and bulk sequencing data revealed HLA-dependent cellular distribution patterns that provide insights into downstream immune effector mechanisms, critical upstream pathways remain unresolved. It remains unclear whether TiA’s pathogenesis is similar to other HLA-linked drug adverse reactions that are induced by HLA-TCR interactions. Most importantly, the TCR-recognized antigens driving TiA remain unknown, limiting direct validation of TCR-antigen interactions. Lastly, the limited number of cells obtained from single-cell sequencing was insufficient for comprehensive statistical analysis. Future studies integrating larger sample sizes, increased sequencing depth, expanded single-cell datasets, and critically, identification of TiA-relevant antigens, will be essential to elucidate the complete pathogenic pathway from antigen recognition to agranulocytosis. Such mechanistic understanding would facilitate the development of predictive biomarkers and targeted therapeutic interventions for TiA.

## CONCLUSION

According to our previous genome-wide association study, *HLA-B*38:02* and *HLA-DRB1*08:03* were identified as independent genetic susceptibility factors for TiA. HLA genotyping of our cohort validated this association, demonstrating significant enrichment of these risk alleles in TiA patients compared to GD subjects. At the repertoire level, immune repertoire profiling showed that global repertoire composition remained comparable between TiA and GD patients, including diversity, V/J gene usage, and V-J pairing pattern. In contrast, clonotype-level analysis uncovered several TCR and BCR risk clonotypes that were significantly enriched among TiA-acute patients, highlighting their specific roles in TiA pathogenesis. Furthermore, a subset of TCR risk clonotypes with HLA-linked patterns exhibited distinct motif patterns between *HLA-B*38:02* and *HLA-DRB1*08:03* groups. Integration of bulk and single-cell immune repertoire with transcriptome sequencing demonstrated that these TiA-associated risk clonotypes were differentially distributed across T cell subsets in an HLA-dependent manner.

Overall, our study identified TiA-associated clonal signatures and provided novel insights into the immunogenetic and cellular features associated with TiA. These findings highlight the potential of these clonal signatures as predictive biomarkers for risk stratification and as therapeutic targets in TiA.

## MATERIALS AND METHODS

### Study Populations

The overall study population and sample selection strategy were illustrated in Figure 1. Three cohorts designated as ViRep, iRep, and iRepC were collected from National Taiwan University Hospital (NTUH), including patients with diagnosed Graves’ disease who developed TiA, patients with Graves’ disease without TiA, and Controls. Since previous studies have demonstrated no significant difference in TiA incidence between patients receiving PTU and those receiving MMI^43^, drug type was not considered as a stratification factor in our cohort design. Based on the previously identified two TiA-related HLA risk alleles^13^, subjects from these cohorts with defined HLA alleles status (groups I-IV) were enrolled in this study: (I) subjects with only *HLA-B*38:02* (abbreviated as B*38:02 only), (II) those possessing only *HLA-DRB1*08:03* (abbreviated as DRB1*08:03 only), (III) subjects harboring both risk alleles (abbreviated as Both), (IV) those lacking both alleles (abbreviated as Without) (Table S1). Each group comprised TiA patients, GD patients, and Controls. Longitudinal samples were also obtained from selected individuals during the acute phase (at the time of initial diagnosis) and the steady phase (typically 0.5-1 year after diagnosis). In total, 82 samples from 53 patients (8 TiA, 21 GD, and 24 healthy controls) were included in immune repertoire sequencing.

### HLA Genotyping

HLA genotyping was performed on five cohorts including the three cohorts mentioned above and additional GD and VTIA cohorts, totaling 65 TiA patients, 167 GD patients, and 125 normal controls. Genomic DNA from these samples was subjected to library preparation, probe capture-based enrichment, and sequencing as previously described^21^.

### Bulk Immune Repertoire Sequencing

The overall experimental workflow was illustrated in Figure 1A. RNA from PBMCs of participants were extracted using 1ml TRIzol reagents (Invitrogen, Carlsbad, USA), followed by preparation of cDNA libraries using the human NEBNext Immune Sequencing Kit (New England Biolabs, Massachusetts, USA). The library construction employed reverse transcription (RT) with oligo (dT) primers, followed by template switching with unique molecular identifiers (UMIs) incorporation for accurate quantification. PCR amplification was performed using P7 index primers and BCR/TCR-specific primers targeting the constant region (C-region primers) to capture the complete V(D)J gene segments, followed by the addition of P5 index primers to complete the library preparation for paired-end sequencing. Additionally, a schematic representation of the paired-end 2×300 sequencing library structure, showing the positions and length of UMIs, template switch (TS) sequences, and C-region primers, is shown in Figure 3. High-throughput next-generation sequencing (NGS) was performed on the NextSeq 2000 platform (Illumina, California, USA) to sequence the V(D)J junction of immune repertoire.

Subsequently, the computational algorithm TRUST4^44^ was applied to reconstruct and defined the rearranged CDR3 of T/BCR repertoires according to the ImMunoGeneTics information (IMGT) database (www.imgt.org). TRUST4 output files contained CDR3 clonotypes (abbreviated as clonotypes), defined as groups of sequences sharing identical CDR3 nucleotide sequences, along with their corresponding V(D)J gene usage, CDR3 nucleotide and amino acid sequences, and read counts, which were subjected to post-analysis.

### Bulk Immune Repertoire Post-analysis

Following AIRR community guidelines, analysis typically focuses on diversity, V/J gene usages, VJ associations, and shared CDR3 clonotype to understand immune repertoire characteristics and underlying immune-mediated pathophysiology^45^.

To improve analytical resolution and avoid the interference from rare clonotypes, clonotypes with only one count were excluded from subsequent analyses. All CDR3 sequences were initially categorized by different TCR (TRA/TRB) and BCR (IGH/IGK/IGL) regions. Overall repertoire diversity was estimated through three indices: relative D50, Shannon entropy, Simpson’s dominance. The relative D50, a measure of repertoire richness, was defined as:

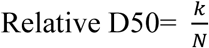

where *k* denotes the minimal number of top-ranked clonotypes whose cumulative frequency exceeds 50% and N is the total number of unique clonotypes in the repertoire. Lower relative D50 values indicate repertoires dominated by fewer high-abundance clonotypes. To quantify repertoire evenness, Shannon entropy was computed as:

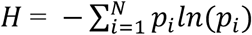

where *p_i_* represents the relative frequency of clonotype *i* and *N* is the total number of clonotypes. Simpson’s dominance was calculated as:

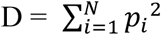

with higher value indicating greater dominance and lower diversity.

V/J gene usage frequency was calculated by dividing individual V or J gene counts by the total count within each region. Similarly, V-J pairing frequency was determined by dividing specific V-J combination counts by the total count within the corresponding region. Cosine similarity between groups was calculated using the standard cosine similarity equation, where each group was represented as a vector of mean V-J pairing frequencies across all samples within that group.

To identify disease-associated clonotypes, we employed a dual analytical approach combining frequency-based and incidence-based analyses. Frequency of CDR3 clonotypes was evaluated by dividing counts of each unique CDR3 clonotype by the total count within the corresponding region. For the frequency-based analysis of TiA-associated and GD-associated clonotype identification, clonotypes were identified using a significance threshold of *p* < 0.05 (Wilcoxon rank-sum test) (*p* < 0.05, Wilcoxon rank-sum test). For the incidence-based analysis, GD-associated clonotypes were identified using Fisher’s exact test (*p* < 0.05), whereas TiA-associated clonotypes were instead defined as those present in more than two TiA-acute subjects (n > 2) and absent in all GD-acute patients, given that the limited TiA-acute sample size (n = 8) and imbalance with GD-acute (n = 21) precluded the use of Fisher’s exact test.

TCR CDR3 sequence similarity was evaluated using *tcrdist3*^26^, while BCR CDR3 similarity was assessed using BLOSUM62-based distance metrics adapted from *tcrdist3*. Hierarchical clustering was applied at a distance threshold of 16 to identify convergent clonotypes. Sequence motifs were visualized using position weight matrices derived from multiple sequence alignment.

### Single-cell Immune Repertoire Sequencing with Whole Transcriptomic Profiling

To simultaneously profile the whole transcriptome and immune repertoire at single-cell resolution, we utilized the Evercode™ TCR (Parse Biosciences). Single-cell suspensions were fixed, permeabilized, and subjected to split-pool combinatorial barcoding following the manufacturer’s standard instructions.

TCR and whole-transcriptome libraries were sequenced on the NextSeq and NovaSeq X Plus Platforms (Illumina, California, USA), respectively. Raw data were processed using Seurat (v 5.4.0)^46^ for quality control, normalization, dimensionality reduction, and clustering. Cell type annotation was performed using Azimuth with the default human PBMC reference^47^. Following annotation, B cell clusters exhibiting concurrent TCR transcript were identified as putative B-T doublets. Given that these clusters demonstrated a mean nFeature_RNA exceeding 4,000 genes per cell, they were manually excluded from all downstream analyses.

This approach enabled the precise capture of paired TCR α and β chain sequences along with full transcriptomic data, facilitating the comprehensive characterization of T cell subsets and clonotype tracking.

### *In Silico* Structure Prediction and Binding Affinity Assessment of BCR Clonotypes

The crystal structure of thyroid-stimulating hormone receptor (TSHR) bound to thyroid-stimulating autoantibody was obtained from the Protein Data Bank (PDB accession code: 3G04)^24^ and used as the reference structure. Risk and non-risk BCR sequences (IGK) were selected from GD patients during the acute phase compared to normal control based on our repertoire analysis. The structures of selected IGK paired with IGH derived from the reference structure were generated using AlphaFold 3^48^. Antibody-TSHR docking was performed using ClusPro, a widely-used protein-protein docking tool^28–32^. PyMOL (PyMOL Molecular Graphics System, v 3.1.4.1, Schrodinger LLC, New York, NY) was used to visualize the docking results and evaluate epitope coverage by comparing the antibody-TSHR interface with the reference structure. Binding energies were estimated using PRODIGY^33, 34^.

### Statistical Analysis

Statistical differences between the two groups are explained in the respective figure legends. R (v 4.3.1) and Python (v 3.12.4) were used for advanced statistical data analysis and visualization.

## DECLARATIONS

### Ethics approval and consent to participate

The study involved human participants and was conducted in accordance with the Declaration of Helsinki. The Research Ethics Committee of National Taiwan University Hospital gave ethical approval for this work (Approval No. 201611020RIND). Written informed consent was obtained from all participants.

### Consent for publication

Not applicable.

### Availability of data and materials

The datasets used and/or analyzed during the current study are available from the corresponding author on reasonable request.

### Author’s contributions

Conceptualization, H.Y.H., L.S.K., J.S.H., C.C.Y., H.C.L., Y.Y.C., and C.P.L.; Methodology, H.Y.H., L.S.K., and C.I.H.; Formal analysis, H.Y.H., L.S.K., Y.Y.H., and C.J.L.; Data curation, H.Y.H., L.S.K., Y.Y.H., and C.I.H.; Resources, C.S.C., L.P.S., L.C.H., C.W.Y., W.W.C., L.J.Y., W.C.Y., C.C.Y., Y.W.S., H.C.J., S.S.R., and C.P.L.; Writing – original draft, H.Y.H., and L.S.K.; Writing – review and editing, all authors. All authors read and approved the final manuscript.

## Acknowledgements

We thank to National Center for High-performance Computing (NCHC) for providing computational and storage resources.

