## Supplementary Data for "Integrative Genomic and Immune Repertoire Profiling Identifies Clonal Signatures Linked to Antithyroid Drug-Induced Agranulocytosis"

**SUPPLEMENTARY FIGURES**

Figure S1. Tree map visualization of clonal distribution in individual samples.

Figure S2. V/J gene usage patterns across disease phenotypes and phases.

Figure S3. V-J pairing patterns among different disease phenotypes.

Figure S4. Structural analysis of GD-associated IGK clonotypes.

Figure S5. Further analyses for TiA-associated TCR risk clonotypes.

Figure S6. Identification of TiA-associated BCR risk clonotypes.

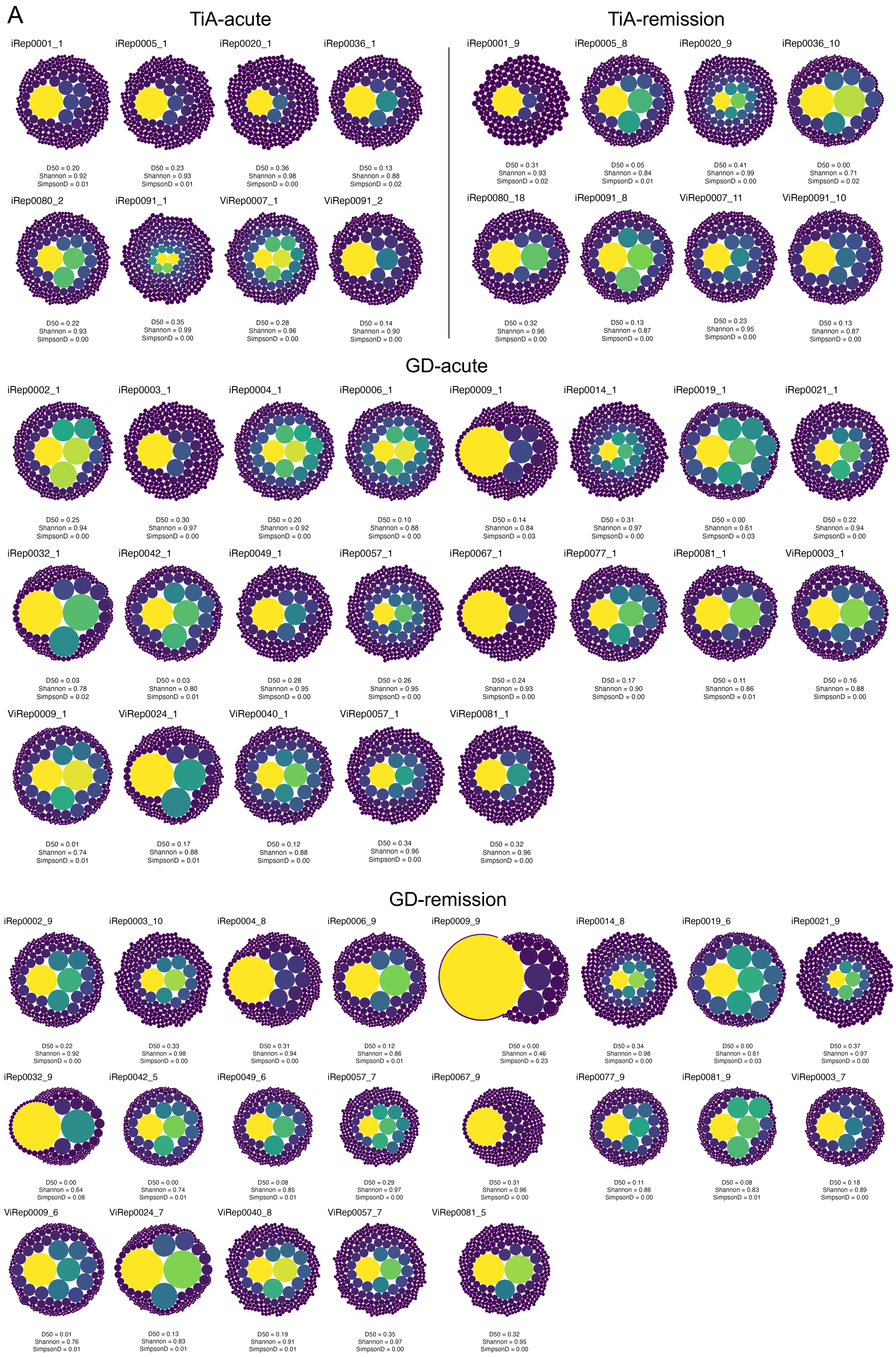
A
TiA-acute
TiA-remission
iRep0001_1
D50 = 0.20
Shannon = 0.92
SimpsonD = 0.01
iRep0005_1
D50 = 0.23
Shannon = 0.93
SimpsonD = 0.01
iRep0020_1
D50 = 0.36
Shannon = 0.98
SimpsonD = 0.00
iRep0036_1
D50 = 0.13
Shannon = 0.88
SimpsonD = 0.02
iRep0001_9
D50 = 0.31
Shannon = 0.93
SimpsonD = 0.02
iRep0005_8
D50 = 0.05
Shannon = 0.84
SimpsonD = 0.01
iRep0020_9
D50 = 0.41
Shannon = 0.99
SimpsonD = 0.00
iRep0036_10
D50 = 0.00
Shannon = 0.71
SimpsonD = 0.02
iRep0080_2
D50 = 0.22
Shannon = 0.93
SimpsonD = 0.00
iRep0091_1
D50 = 0.35
Shannon = 0.99
SimpsonD = 0.00
ViRep0007_1
D50 = 0.28
Shannon = 0.96
SimpsonD = 0.00
ViRep0091_2
D50 = 0.14
Shannon = 0.90
SimpsonD = 0.00
iRep0080_18
D50 = 0.32
Shannon = 0.96
SimpsonD = 0.00
iRep0091_8
D50 = 0.13
Shannon = 0.87
SimpsonD = 0.00
ViRep0007_11
D50 = 0.23
Shannon = 0.95
SimpsonD = 0.00
ViRep0091_10
D50 = 0.13
Shannon = 0.87
SimpsonD = 0.00
GD-acute
iRep0002_1
D50 = 0.25
Shannon = 0.94
SimpsonD = 0.00
iRep0003_1
D50 = 0.30
Shannon = 0.97
SimpsonD = 0.00
iRep0004_1
D50 = 0.20
Shannon = 0.92
SimpsonD = 0.00
iRep0006_1
D50 = 0.10
Shannon = 0.88
SimpsonD = 0.00
iRep0009_1
D50 = 0.14
Shannon = 0.84
SimpsonD = 0.03
iRep0014_1
D50 = 0.31
Shannon = 0.97
SimpsonD = 0.00
iRep0019_1
D50 = 0.00
Shannon = 0.61
SimpsonD = 0.03
iRep0021_1
D50 = 0.22
Shannon = 0.94
SimpsonD = 0.00
iRep0032_1
D50 = 0.03
Shannon = 0.78
SimpsonD = 0.02
iRep0042_1
D50 = 0.03
Shannon = 0.80
SimpsonD = 0.01
iRep0049_1
D50 = 0.28
Shannon = 0.95
SimpsonD = 0.00
iRep0057_1
D50 = 0.26
Shannon = 0.95
SimpsonD = 0.00
iRep0067_1
D50 = 0.24
Shannon = 0.93
SimpsonD = 0.00
iRep0077_1
D50 = 0.17
Shannon = 0.90
SimpsonD = 0.00
iRep0081_1
D50 = 0.11
Shannon = 0.86
SimpsonD = 0.01
ViRep0003_1
D50 = 0.16
Shannon = 0.88
SimpsonD = 0.00
ViRep0009_1
D50 = 0.01
Shannon = 0.74
SimpsonD = 0.01
ViRep0024_1
D50 = 0.17
Shannon = 0.88
SimpsonD = 0.01
ViRep0040_1
D50 = 0.12
Shannon = 0.88
SimpsonD = 0.00
ViRep0057_1
D50 = 0.34
Shannon = 0.96
SimpsonD = 0.00
ViRep0081_1
D50 = 0.32
Shannon = 0.96
SimpsonD = 0.00
GD-remission
iRep0002_9
D50 = 0.22
Shannon = 0.92
SimpsonD = 0.00
iRep0003_10
D50 = 0.33
Shannon = 0.98
SimpsonD = 0.00
iRep0004_8
D50 = 0.31
Shannon = 0.94
SimpsonD = 0.00
iRep0006_9
D50 = 0.12
Shannon = 0.86
SimpsonD = 0.01
iRep0009_9
D50 = 0.00
Shannon = 0.46
SimpsonD = 0.23
iRep0014_8
D50 = 0.34
Shannon = 0.98
SimpsonD = 0.00
iRep0019_6
D50 = 0.00
Shannon = 0.61
SimpsonD = 0.03
iRep0021_9
D50 = 0.37
Shannon = 0.97
SimpsonD = 0.00
iRep0032_9
D50 = 0.00
Shannon = 0.64
SimpsonD = 0.08
iRep0042_5
D50 = 0.00
Shannon = 0.74
SimpsonD = 0.01
iRep0049_6
D50 = 0.08
Shannon = 0.85
SimpsonD = 0.01
iRep0057_7
D50 = 0.29
Shannon = 0.97
SimpsonD = 0.00
iRep0067_9
D50 = 0.31
Shannon = 0.96
SimpsonD = 0.00
iRep0077_9
D50 = 0.11
Shannon = 0.86
SimpsonD = 0.00
iRep0081_9
D50 = 0.08
Shannon = 0.83
SimpsonD = 0.01
ViRep0003_7
D50 = 0.18
Shannon = 0.89
SimpsonD = 0.00
ViRep0009_6
D50 = 0.01
Shannon = 0.76
SimpsonD = 0.01
ViRep0024_7
D50 = 0.13
Shannon = 0.83
SimpsonD = 0.01
ViRep0040_8
D50 = 0.19
Shannon = 0.91
SimpsonD = 0.01
ViRep0057_7
D50 = 0.35
Shannon = 0.97
SimpsonD = 0.00
ViRep0081_5
D50 = 0.32
Shannon = 0.95
SimpsonD = 0.00

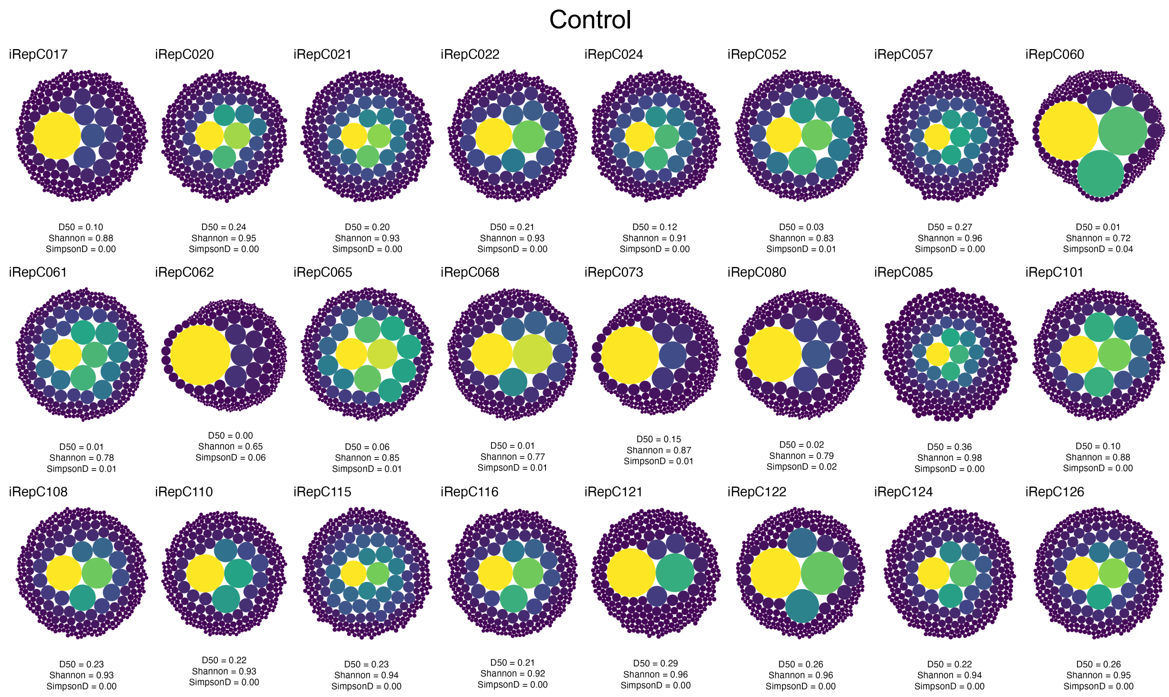

Control
iRepC017
D50 = 0.10
Shannon = 0.88
SimpsonD = 0.00
iRepC020
D50 = 0.24
Shannon = 0.95
SimpsonD = 0.00
iRepC021
D50 = 0.20
Shannon = 0.93
SimpsonD = 0.00
iRepC022
D50 = 0.21
Shannon = 0.93
SimpsonD = 0.00
iRepC024
D50 = 0.12
Shannon = 0.91
SimpsonD = 0.00
iRepC052
D50 = 0.03
Shannon = 0.83
SimpsonD = 0.01
iRepC057
D50 = 0.27
Shannon = 0.96
SimpsonD = 0.00
iRepC060
D50 = 0.01
Shannon = 0.72
SimpsonD = 0.04
iRepC061
D50 = 0.01
Shannon = 0.78
SimpsonD = 0.01
iRepC062
D50 = 0.00
Shannon = 0.65
SimpsonD = 0.06
iRepC065
D50 = 0.06
Shannon = 0.85
SimpsonD = 0.01
iRepC068
D50 = 0.01
Shannon = 0.77
SimpsonD = 0.01
iRepC073
D50 = 0.15
Shannon = 0.87
SimpsonD = 0.01
iRepC080
D50 = 0.02
Shannon = 0.79
SimpsonD = 0.02
iRepC085
D50 = 0.36
Shannon = 0.98
SimpsonD = 0.00
iRepC101
D50 = 0.10
Shannon = 0.88
SimpsonD = 0.00
iRepC108
D50 = 0.23
Shannon = 0.93
SimpsonD = 0.00
iRepC110
D50 = 0.22
Shannon = 0.93
SimpsonD = 0.00
iRepC115
D50 = 0.23
Shannon = 0.94
SimpsonD = 0.00
iRepC116
D50 = 0.21
Shannon = 0.92
SimpsonD = 0.00
iRepC121
D50 = 0.29
Shannon = 0.96
SimpsonD = 0.00
iRepC122
D50 = 0.26
Shannon = 0.96
SimpsonD = 0.00
iRepC124
D50 = 0.22
Shannon = 0.94
SimpsonD = 0.00
iRepC126
D50 = 0.26
Shannon = 0.95
SimpsonD = 0.00

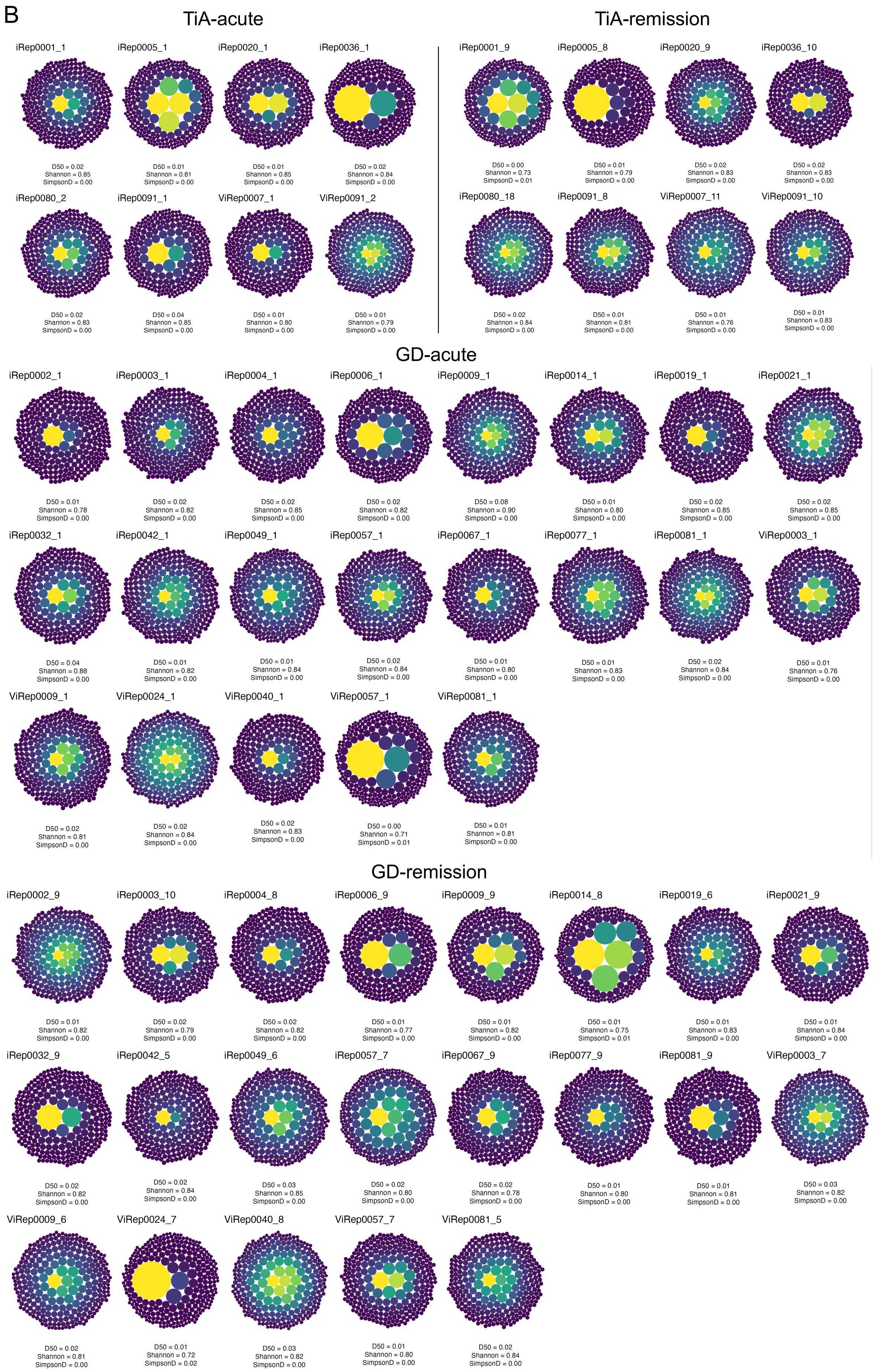
B
TiA-acute
TiA-remission
iRep0001_1
D50 = 0.02
Shannon = 0.85
SimpsonD = 0.00
iRep0005_1
D50 = 0.01
Shannon = 0.81
SimpsonD = 0.00
iRep0020_1
D50 = 0.01
Shannon = 0.85
SimpsonD = 0.00
iRep0036_1
D50 = 0.02
Shannon = 0.84
SimpsonD = 0.00
iRep0001_9
D50 = 0.00
Shannon = 0.73
SimpsonD = 0.01
iRep0005_8
D50 = 0.01
Shannon = 0.79
SimpsonD = 0.00
iRep0020_9
D50 = 0.02
Shannon = 0.83
SimpsonD = 0.00
iRep0036_10
D50 = 0.02
Shannon = 0.83
SimpsonD = 0.00
iRep0080_2
D50 = 0.02
Shannon = 0.83
SimpsonD = 0.00
iRep0091_1
D50 = 0.04
Shannon = 0.85
SimpsonD = 0.00
ViRep0007_1
D50 = 0.01
Shannon = 0.80
SimpsonD = 0.00
ViRep0091_2
D50 = 0.01
Shannon = 0.79
SimpsonD = 0.00
iRep0080_18
D50 = 0.02
Shannon = 0.84
SimpsonD = 0.00
iRep0091_8
D50 = 0.01
Shannon = 0.81
SimpsonD = 0.00
ViRep0007_11
D50 = 0.01
Shannon = 0.76
SimpsonD = 0.00
ViRep0091_10
D50 = 0.01
Shannon = 0.83
SimpsonD = 0.00
GD-acute
iRep0002_1
D50 = 0.01
Shannon = 0.78
SimpsonD = 0.00
iRep0003_1
D50 = 0.02
Shannon = 0.82
SimpsonD = 0.00
iRep0004_1
D50 = 0.02
Shannon = 0.85
SimpsonD = 0.00
iRep0006_1
D50 = 0.02
Shannon = 0.82
SimpsonD = 0.00
iRep0009_1
D50 = 0.08
Shannon = 0.90
SimpsonD = 0.00
iRep0014_1
D50 = 0.01
Shannon = 0.80
SimpsonD = 0.00
iRep0019_1
D50 = 0.02
Shannon = 0.85
SimpsonD = 0.00
iRep0021_1
D50 = 0.02
Shannon = 0.85
SimpsonD = 0.00
iRep0032_1
D50 = 0.04
Shannon = 0.88
SimpsonD = 0.00
iRep0042_1
D50 = 0.01
Shannon = 0.82
SimpsonD = 0.00
iRep0049_1
D50 = 0.01
Shannon = 0.84
SimpsonD = 0.00
iRep0057_1
D50 = 0.02
Shannon = 0.84
SimpsonD = 0.00
iRep0067_1
D50 = 0.01
Shannon = 0.80
SimpsonD = 0.00
iRep0077_1
D50 = 0.01
Shannon = 0.83
SimpsonD = 0.00
iRep0081_1
D50 = 0.02
Shannon = 0.84
SimpsonD = 0.00
ViRep0003_1
D50 = 0.01
Shannon = 0.76
SimpsonD = 0.00
ViRep0009_1
D50 = 0.02
Shannon = 0.81
SimpsonD = 0.00
ViRep0024_1
D50 = 0.02
Shannon = 0.84
SimpsonD = 0.00
ViRep0040_1
D50 = 0.02
Shannon = 0.83
SimpsonD = 0.00
ViRep0057_1
D50 = 0.00
Shannon = 0.71
SimpsonD = 0.01
ViRep0081_1
D50 = 0.01
Shannon = 0.81
SimpsonD = 0.00
GD-remission
iRep0002_9
D50 = 0.01
Shannon = 0.82
SimpsonD = 0.00
iRep0003_10
D50 = 0.02
Shannon = 0.79
SimpsonD = 0.00
iRep0004_8
D50 = 0.02
Shannon = 0.82
SimpsonD = 0.00
iRep0006_9
D50 = 0.01
Shannon = 0.77
SimpsonD = 0.00
iRep0009_9
D50 = 0.01
Shannon = 0.82
SimpsonD = 0.00
iRep0014_8
D50 = 0.01
Shannon = 0.75
SimpsonD = 0.01
iRep0019_6
D50 = 0.01
Shannon = 0.83
SimpsonD = 0.00
iRep0021_9
D50 = 0.01
Shannon = 0.84
SimpsonD = 0.00
iRep0032_9
D50 = 0.02
Shannon = 0.82
SimpsonD = 0.00
iRep0042_5
D50 = 0.02
Shannon = 0.84
SimpsonD = 0.00
iRep0049_6
D50 = 0.03
Shannon = 0.85
SimpsonD = 0.00
iRep0057_7
D50 = 0.02
Shannon = 0.80
SimpsonD = 0.00
iRep0067_9
D50 = 0.02
Shannon = 0.78
SimpsonD = 0.00
iRep0077_9
D50 = 0.01
Shannon = 0.80
SimpsonD = 0.00
iRep0081_9
D50 = 0.01
Shannon = 0.81
SimpsonD = 0.00
ViRep0003_7
D50 = 0.03
Shannon = 0.82
SimpsonD = 0.00
ViRep0009_6
D50 = 0.02
Shannon = 0.81
SimpsonD = 0.00
ViRep0024_7
D50 = 0.01
Shannon = 0.72
SimpsonD = 0.02
ViRep0040_8
D50 = 0.03
Shannon = 0.82
SimpsonD = 0.00
ViRep0057_7
D50 = 0.01
Shannon = 0.80
SimpsonD = 0.00
ViRep0081_5
D50 = 0.02
Shannon = 0.84
SimpsonD = 0.00

1

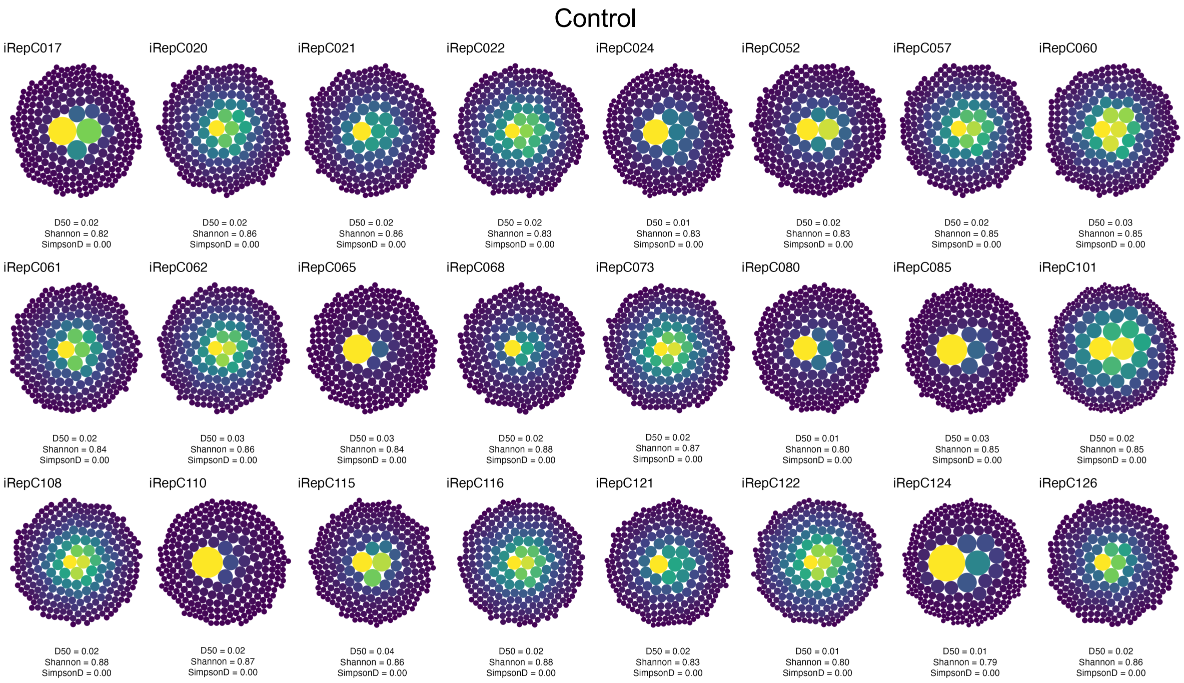


**Figure S1. Tree map visualization of clonal distribution in individual samples.**

(**A-B**) Tree maps showing the top 250 most abundant TCR (A) and BCR (B) clonotypes in each sample. Each circle represents a unique clonotype, with size proportional to relative clone frequency (clonotype count/total counts). Color intensity (viridis scale) indicates clonotype frequency, with brighter colors representing higher frequencies. Diversity indices (Shannon entropy, D50 index, and Simpson's dominance) are annotated below each tree map.

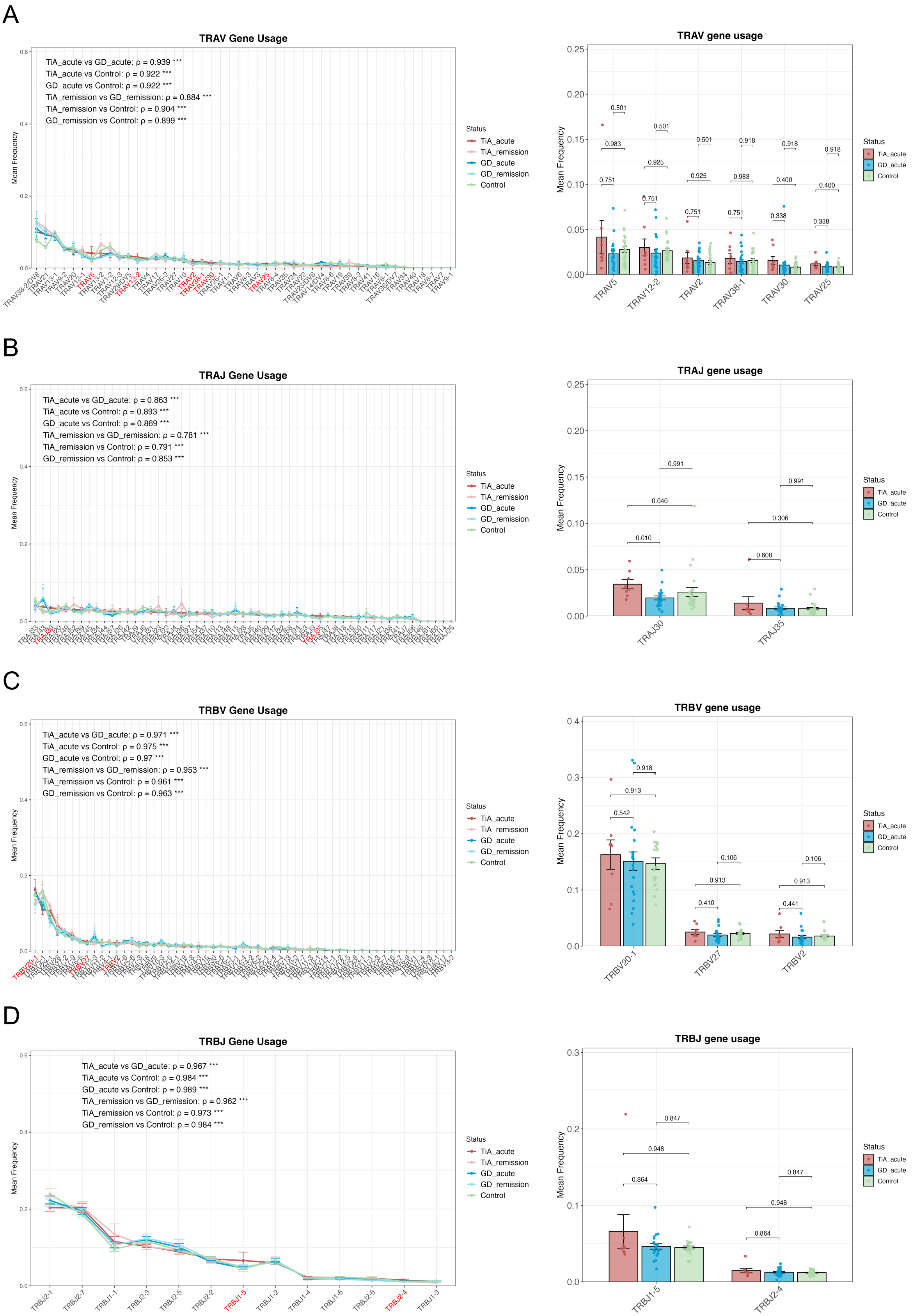
A
TRAV Gene Usage
TiA_acute vs GD_acute: ρ = 0.939 ***
TiA_acute vs Control: ρ = 0.922 ***
GD_acute vs Control: ρ = 0.922 ***
TiA_remission vs GD_remission: ρ = 0.884 ***
TiA_remission vs Control: ρ = 0.904 ***
GD_remission vs Control: ρ = 0.899 ***
Mean Frequency
Status
TiA_acute
TiA_remission
GD_acute
GD_remission
Control
TRAV gene usage
TRAV5
TRAV12-2
TRAV2
TRAV38-1
TRAV30
TRAV25
B
TRAJ Gene Usage
TiA_acute vs GD_acute: ρ = 0.863 ***
TiA_acute vs Control: ρ = 0.893 ***
GD_acute vs Control: ρ = 0.869 ***
TiA_remission vs GD_remission: ρ = 0.781 ***
TiA_remission vs Control: ρ = 0.791 ***
GD_remission vs Control: ρ = 0.853 ***
TRAJ gene usage
TRAJ30
TRAJ35
C
TRBV Gene Usage
TiA_acute vs GD_acute: ρ = 0.971 ***
TiA_acute vs Control: ρ = 0.975 ***
GD_acute vs Control: ρ = 0.97 ***
TiA_remission vs GD_remission: ρ = 0.953 ***
TiA_remission vs Control: ρ = 0.961 ***
GD_remission vs Control: ρ = 0.963 ***
TRBV gene usage
TRBV20-1
TRBV27
TRBV2
D
TRBJ Gene Usage
TiA_acute vs GD_acute: ρ = 0.967 ***
TiA_acute vs Control: ρ = 0.984 ***
GD_acute vs Control: ρ = 0.989 ***
TiA_remission vs GD_remission: ρ = 0.962 ***
TiA_remission vs Control: ρ = 0.973 ***
GD_remission vs Control: ρ = 0.984 ***
TRBJ gene usage
TRBJ1-5
TRBJ2-4

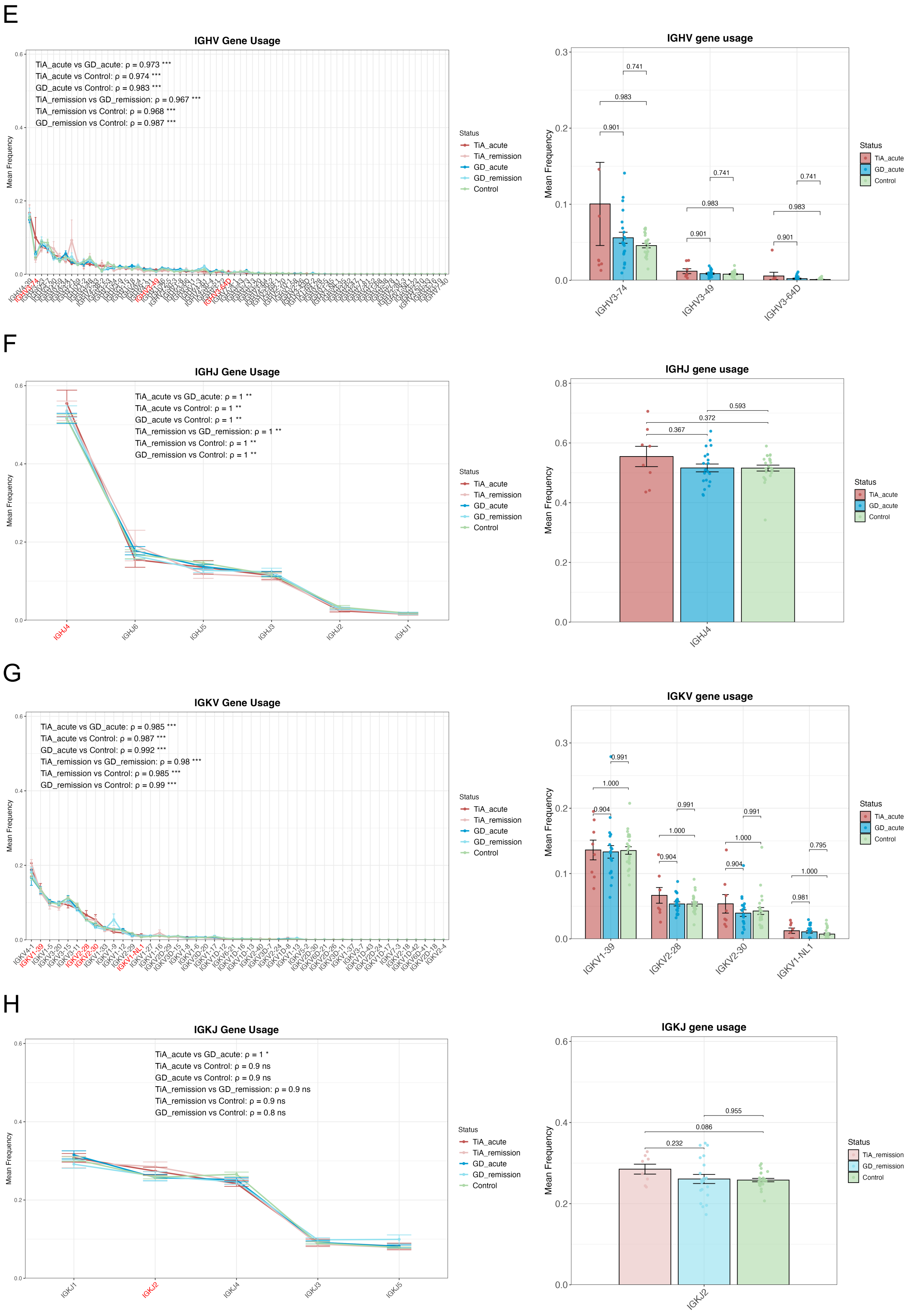
E
IGHV Gene Usage
TiA_acute vs GD_acute: ρ = 0.973 ***
TiA_acute vs Control: ρ = 0.974 ***
GD_acute vs Control: ρ = 0.983 ***
TiA_remission vs GD_remission: ρ = 0.967 ***
TiA_remission vs Control: ρ = 0.968 ***
GD_remission vs Control: ρ = 0.987 ***
Mean Frequency
Status
TiA_acute
TiA_remission
GD_acute
GD_remission
Control
IGHV gene usage
IGHV3-74
IGHV3-49
IGHV3-64D
F
IGHJ Gene Usage
TiA_acute vs GD_acute: ρ = 1 **
TiA_acute vs Control: ρ = 1 **
GD_acute vs Control: ρ = 1 **
TiA_remission vs GD_remission: ρ = 1 **
TiA_remission vs Control: ρ = 1 **
GD_remission vs Control: ρ = 1 **
IGHJ4
IGHJ6
IGHJ5
IGHJ3
IGHJ2
IGHJ1
IGHJ gene usage
G
IGKV Gene Usage
TiA_acute vs GD_acute: ρ = 0.985 ***
TiA_acute vs Control: ρ = 0.987 ***
GD_acute vs Control: ρ = 0.992 ***
TiA_remission vs GD_remission: ρ = 0.98 ***
TiA_remission vs Control: ρ = 0.985 ***
GD_remission vs Control: ρ = 0.99 ***
IGKV gene usage
IGKV1-39
IGKV2-28
IGKV2-30
IGKV1-NL1
H
IGKJ Gene Usage
TiA_acute vs GD_acute: ρ = 1 *
TiA_acute vs Control: ρ = 0.9 ns
GD_acute vs Control: ρ = 0.9 ns
TiA_remission vs GD_remission: ρ = 0.9 ns
TiA_remission vs Control: ρ = 0.9 ns
GD_remission vs Control: ρ = 0.8 ns
IGKJ1
IGKJ2
IGKJ4
IGKJ3
IGKJ5
IGKJ gene usage

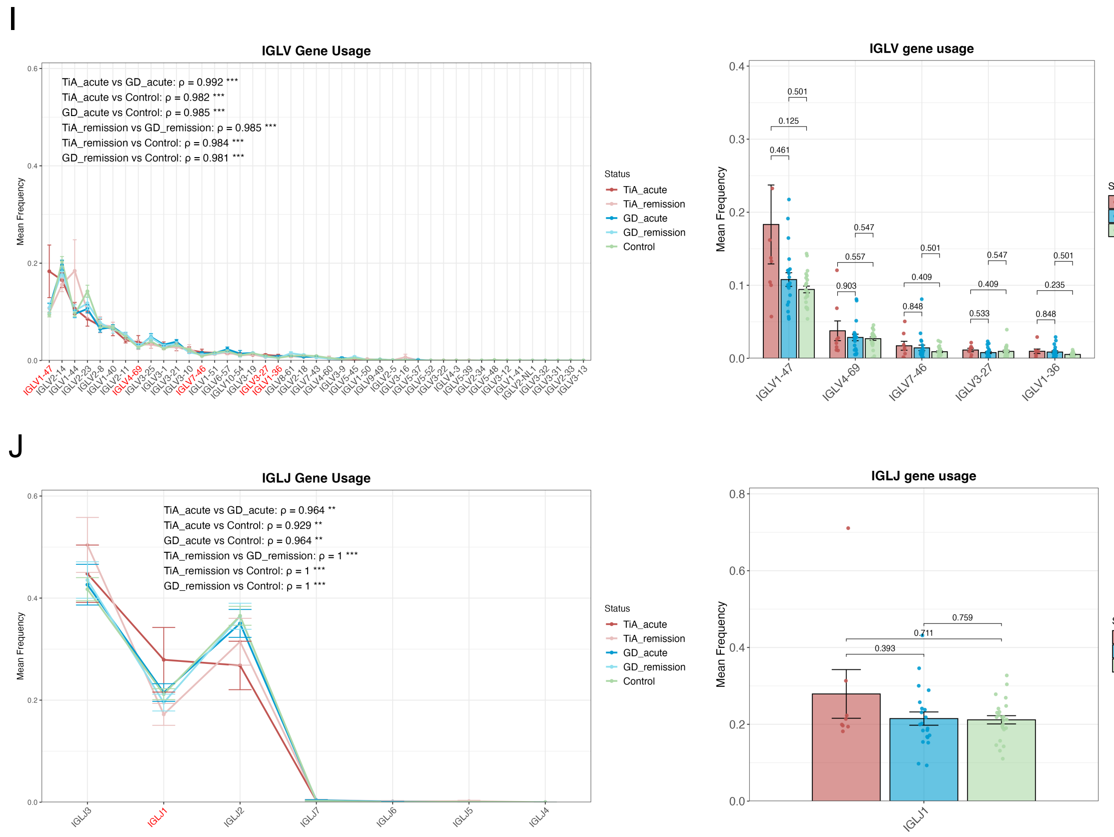


**Figure S2. V/J gene usage patterns across disease phenotypes and phases.**

(**A-J**) Left: TRAV (A), TRAJ (B), TRBV (C), TRBJ (D), IGHV (E), IGHJ (F), IGKV (G), IGKJ (H), IGLV (I), and IGLJ (J) gene usage patterns were shown in line plots (mean ± SD). Spearman's correlation coefficients (ρ) quantify pairwise similarity in gene usage patterns between groups, with higher values indicating greater concordance. Genes enriched in TiA patients (acute or remission phases) are highlighted in red. Statistical significance derived from the Spearman rank correlation test was determined as *$p < 0.05$, **$p < 0.01$, ***$p < 0.001$. Right: Scatter dot plots of TiA-enriched genes across groups (mean ± SD). Statistical significance was assessed using two-sided Wilcoxon rank-sum test with BH correction.

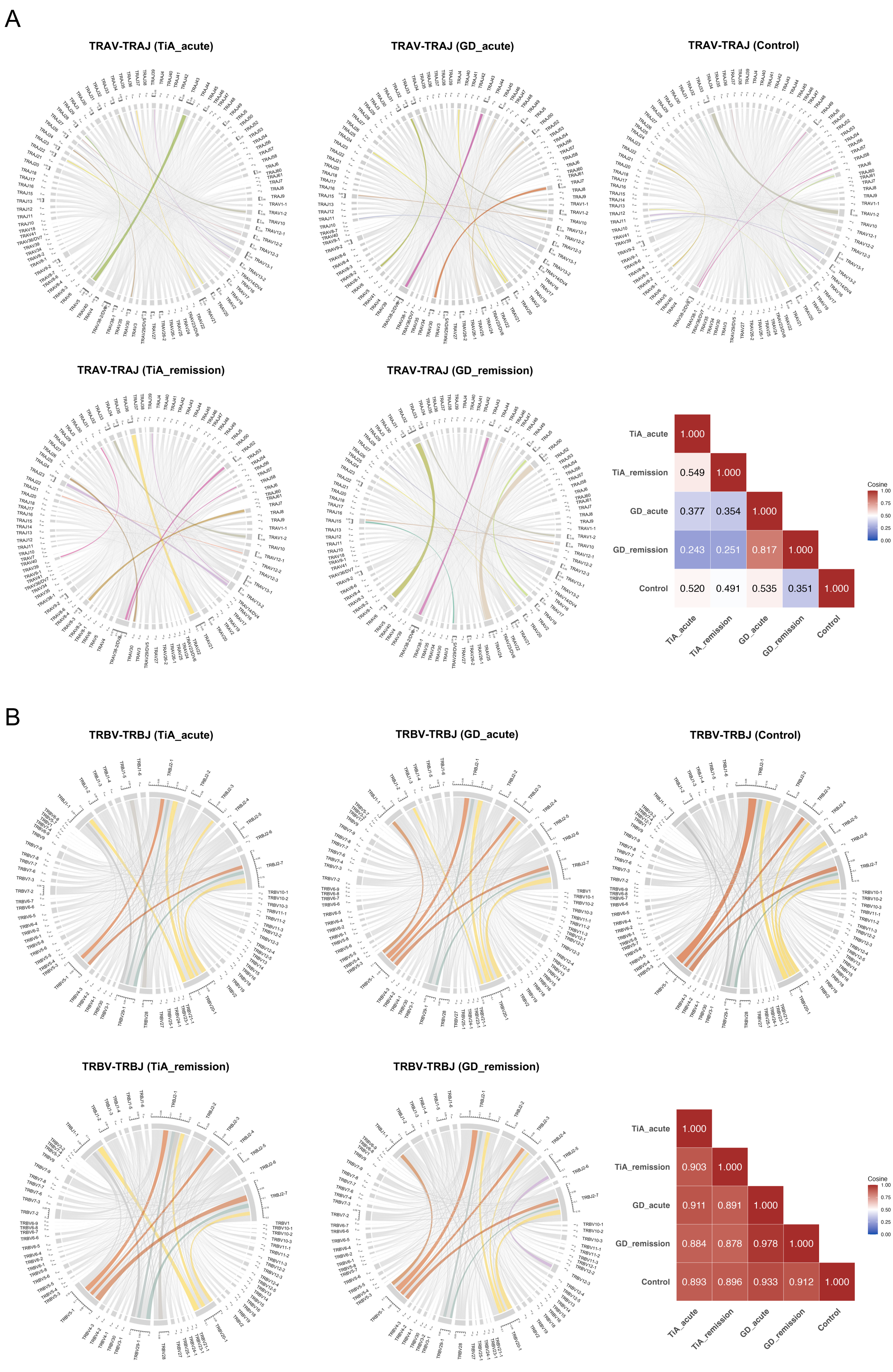
A
TRAV-TRAJ (TiA_acute)
TRAV-TRAJ (GD_acute)
TRAV-TRAJ (Control)
TRAV-TRAJ (TiA_remission)
TRAV-TRAJ (GD_remission)
TiA_acute 1.000
TiA_remission 0.549 1.000
GD_acute 0.377 0.354 1.000
GD_remission 0.243 0.251 0.817 1.000
Control 0.520 0.491 0.535 0.351 1.000
TiA_acute TiA_remission GD_acute GD_remission Control
Cosine
1.00
0.75
0.50
0.25
0.00
B
TRBV-TRBJ (TiA_acute)
TRBV-TRBJ (GD_acute)
TRBV-TRBJ (Control)
TRBV-TRBJ (TiA_remission)
TRBV-TRBJ (GD_remission)
TiA_acute 1.000
TiA_remission 0.903 1.000
GD_acute 0.911 0.891 1.000
GD_remission 0.884 0.878 0.978 1.000
Control 0.893 0.896 0.933 0.912 1.000
TiA_acute TiA_remission GD_acute GD_remission Control
Cosine
1.00
0.75
0.50
0.25
0.00

C

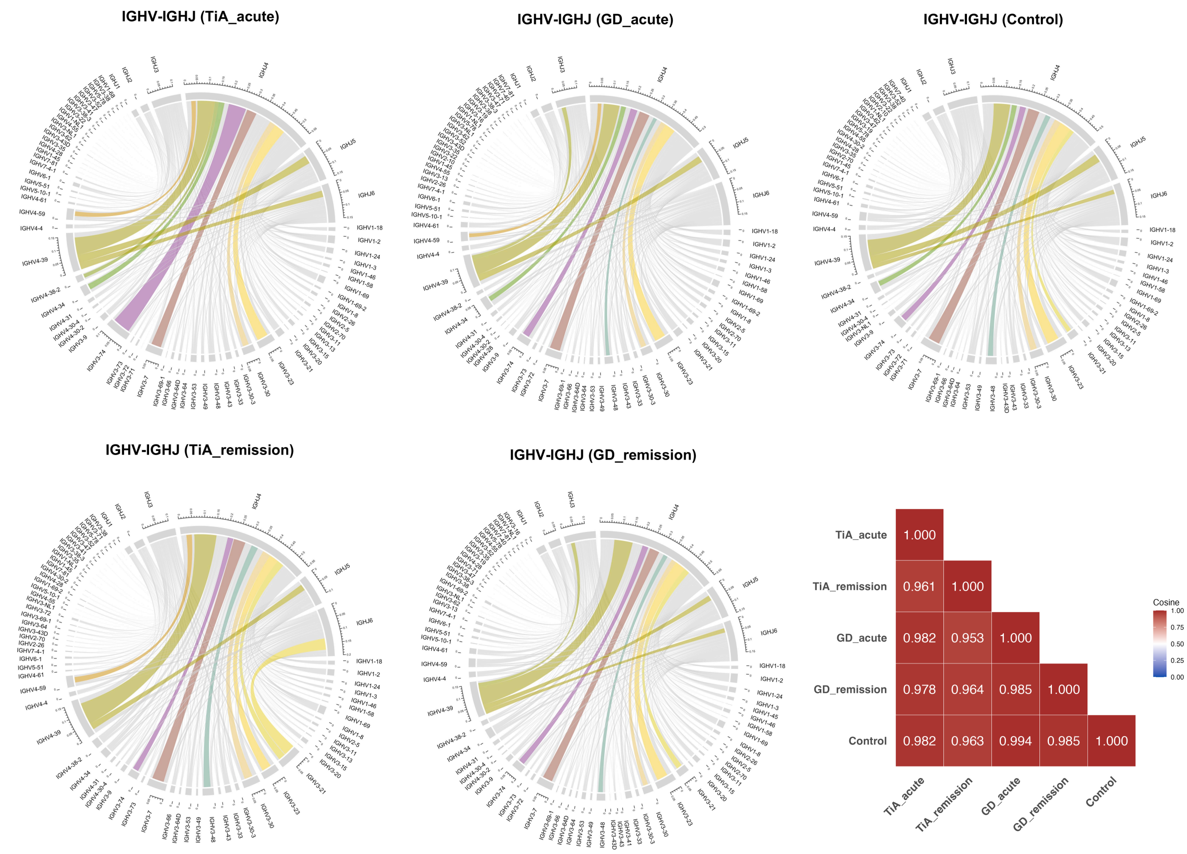

IGHV-IGHJ (TiA_acute)
IGHV-IGHJ (GD_acute)
IGHV-IGHJ (Control)
IGHV-IGHJ (TiA_remission)
IGHV-IGHJ (GD_remission)
TiA_acute 1.000
TiA_remission 0.961 1.000
GD_acute 0.982 0.953 1.000
GD_remission 0.978 0.964 0.985 1.000
Control 0.982 0.963 0.994 0.985 1.000
TiA_acute
TiA_remission
GD_acute
GD_remission
Control
Cosine
1.00
0.75
0.50
0.25
0.00


D

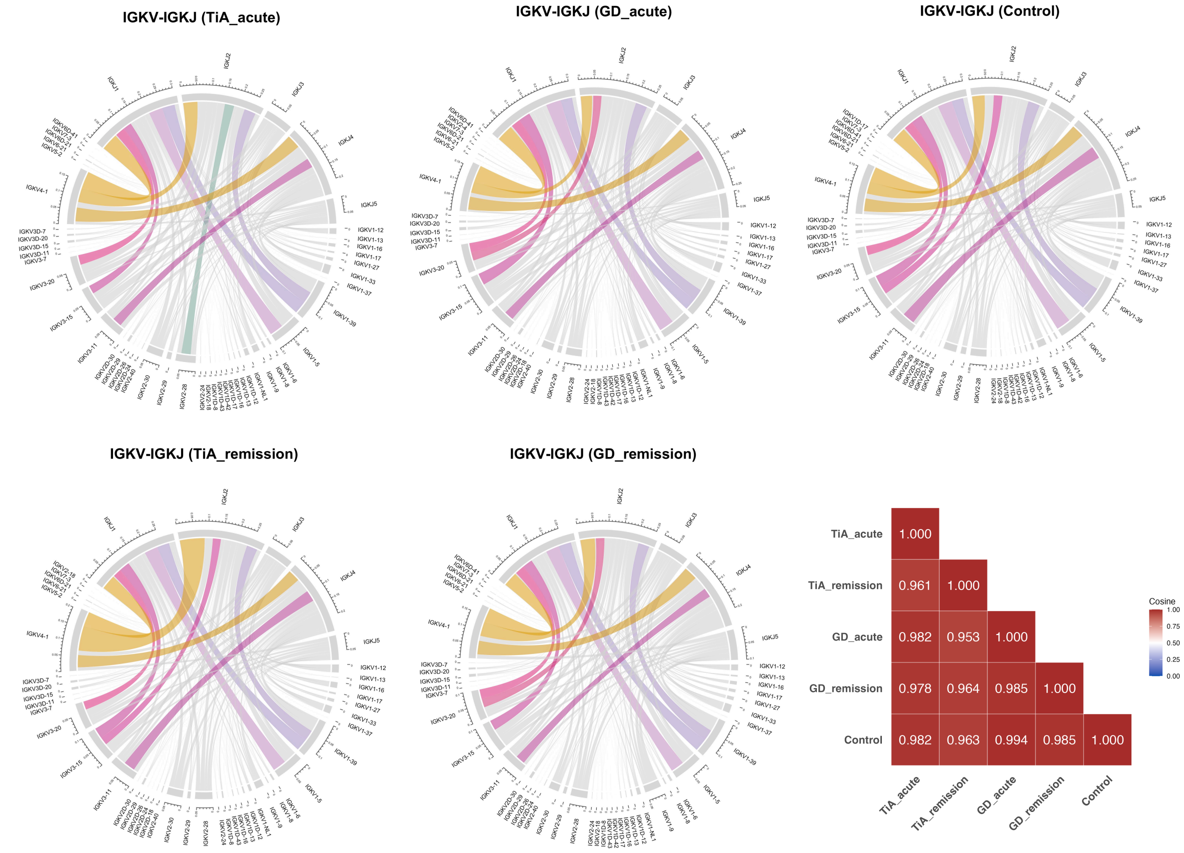

IGKV-IGKJ (TiA_acute)
IGKV-IGKJ (GD_acute)
IGKV-IGKJ (Control)
IGKV-IGKJ (TiA_remission)
IGKV-IGKJ (GD_remission)
TiA_acute 1.000
TiA_remission 0.961 1.000
GD_acute 0.982 0.953 1.000
GD_remission 0.978 0.964 0.985 1.000
Control 0.982 0.963 0.994 0.985 1.000
TiA_acute
TiA_remission
GD_acute
GD_remission
Control
Cosine
1.00
0.75
0.50
0.25
0.00

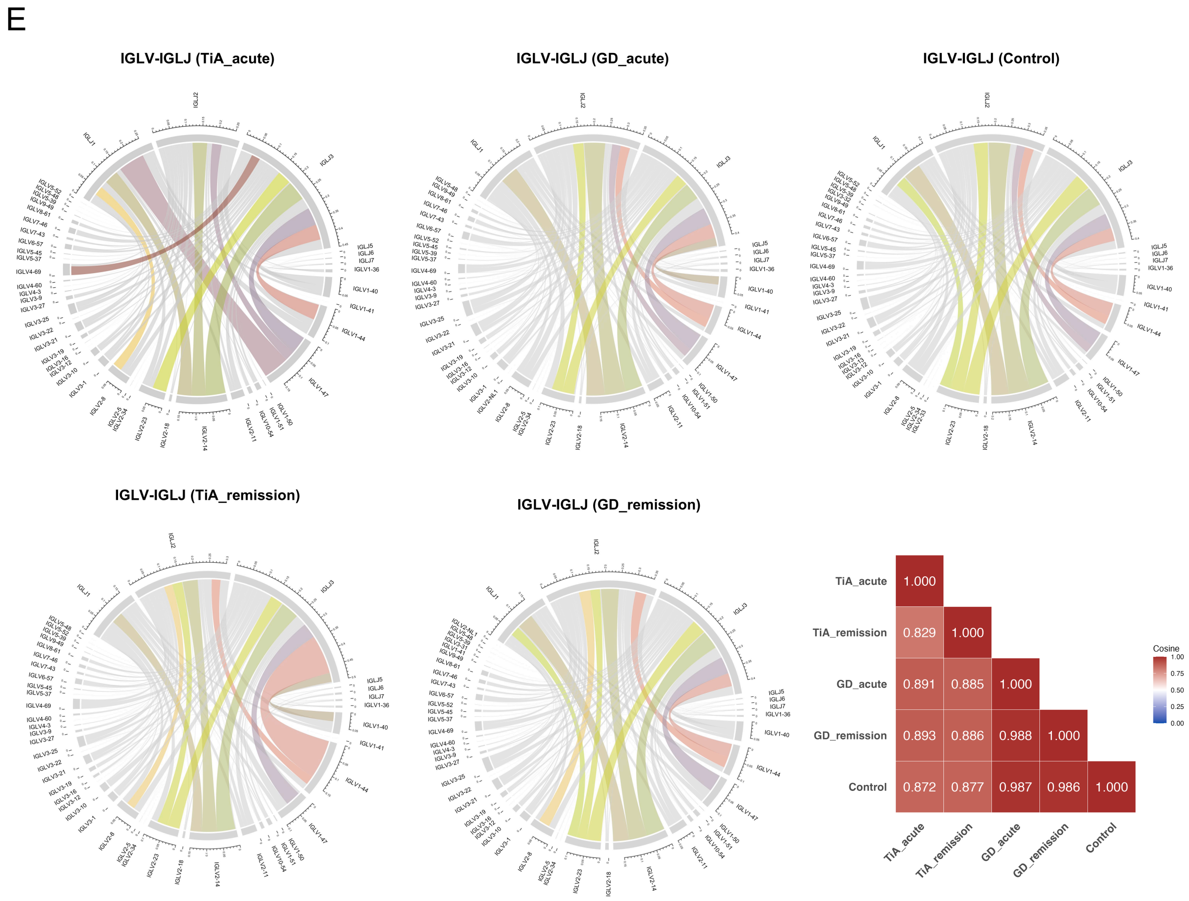


**Figure S3. V-J pairing patterns among different disease phenotypes.**

(**A-E**) Circos plots displaying the frequency of *TRAV-TRAJ* (**A**), *TRBV-TRBJ* (**B**), *IGHV-IGHJ* (**C**), *IGKV-IGKJ* (**D**), and *IGLV-IGLJ* (**E**) pairings across the five groups. The top 10 V-J pairs are color-coded, with identical V-J pairs sharing the same color within each receptor locus. Heatmaps show pairwise cosine similarity between groups, ranging from 0 (no overlap) to 1 (complete overlap).

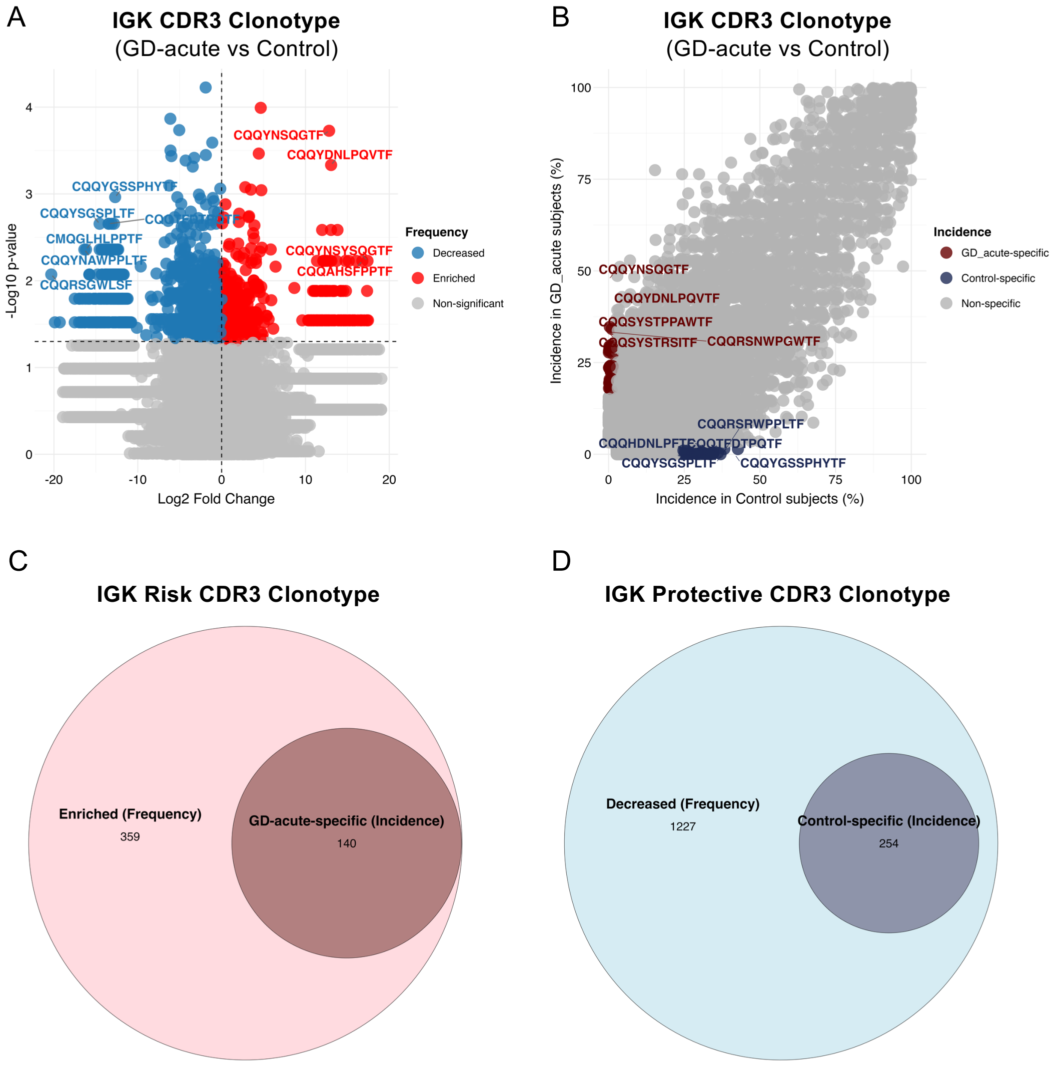
A
IGK CDR3 Clonotype
(GD-acute vs Control)
-Log10 p-value
Log2 Fold Change
CQQYNSQGTF
CQQYDNLPQVTF
CQQYNSYSQGTF
CQQAHSFPPTF
CQQYGSSPHYTF
CQQYSGSPLTF
CMQGLHLPPTF
CQQYNAWPPLTF
CQQRSGWLSF
Frequency
Decreased
Enriched
Non-significant
B
IGK CDR3 Clonotype
(GD-acute vs Control)
Incidence in GD_acute subjects (%)
Incidence in Control subjects (%)
CQQYNSQGTF
CQQYDNLPQVTF
CQQSYSTPPAWTF
CQQSYSTRSITF
CQQRSNWPGWTF
CQQRSRWPPLTF
CQQHDNLPFTF
CQQTFDTPQTF
CQQYSGSPLTF
CQQYGSSPHYTF
Incidence
GD_acute-specific
Control-specific
Non-specific
C
IGK Risk CDR3 Clonotype
Enriched (Frequency)
359
GD-acute-specific (Incidence)
140
D
IGK Protective CDR3 Clonotype
Decreased (Frequency)
1227
Control-specific (Incidence)
254

1

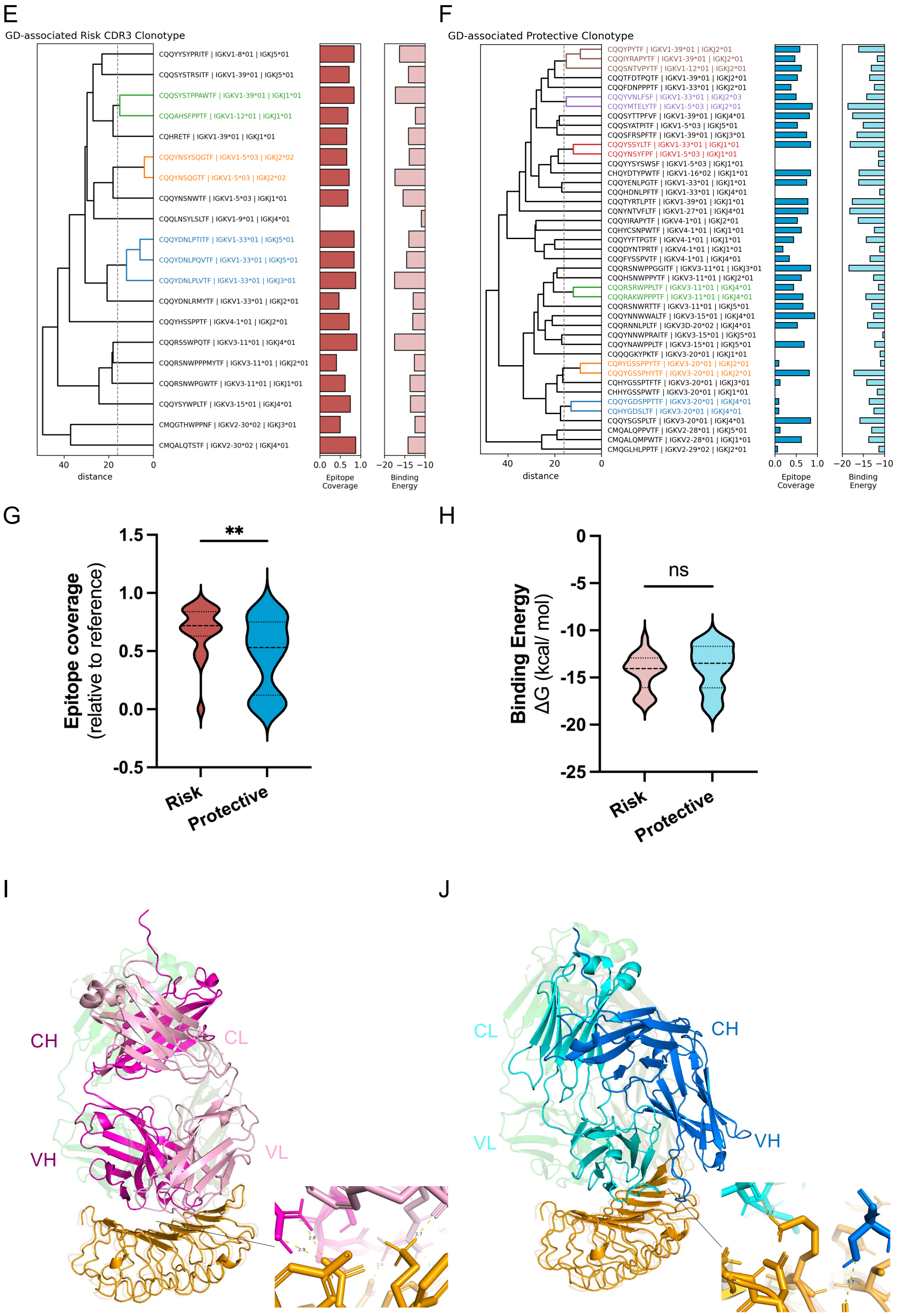


**Figure S4. Structural analysis of GD-associated IGK clonotypes.**

**(A)** Volcano plots showing IGK CDR3 clonotypes with $Log_2$ FC versus $-Log_{10}$ (*p*-value) in GD-acute patients compared to normal control subjects. Statistical significance was calculated using Wilcoxon rank-sum test. Red dots indicate significant enriched clonotypes ($p < 0.05$ and $Log_2$ FC > 0); blue dots indicate significant decreased clonotypes ($p < 0.05$ and $Log_2$ FC < 0); and gray dots represent non-significant clonotypes. CDR3 sequences of the most significant clonotypes are labeled. (**B**) Bubble plots illustrating the clonotype incidence in GD-acute patients versus normal controls. Dark red dots indicate clonotypes present exclusively in GD-acute patients and absent in all control subjects.; dark blue dots indicate clonotypes present exclusively in control subjects and absent in all GD-acute patients. Gray dots represent clonotypes without significant incidence differences. (**C-D**) Venn diagrams showing the overlap between frequency-based and incidence-based GD-associated risk clonotypes (C; enriched in GD-acute or exclusive to GD-acute patients) and protective clonotypes (D; decreased in GD-acute or exclusive to control subjects). Numbers indicate clonotype counts in each category. (**E-F**) Hierarchical clustering of CDR3 sequences based on sequence similarity for GD-associated risk clonotypes (E) and protective clonotypes (F). Clonotypes with distance < 16 were clustered together and highlighted in colors. Heatmaps show epitope coverage (left, relative to reference structure) and binding energy (right, $\Delta G$ in kcal/mol) for each clonotype after antibody-TSHR docking. (**G-H**) Comparison of epitope coverage (G) and binding energy (H) between risk and protective clonotypes. Statistical significance was assessed using the Wilcoxon rank-sum test (**$p < 0.01$; ns, not significant). (**I-J**) Structural comparison of the top-ranked risk clonotype (CQQRSSWPQTF; *IGKV3-11*01*, *IGKJ4*01*) (I) and protective clonotype (CQQSNTVPYTF; *IGKV1-12*01*, *IGKJ2*01*) (J) with the reference structure. Colors indicate heavy chain (CH), light chain (CL), variable regions (VH, VL), and TSHR (orange) for risk (magenta tones), protective (blue tones), and reference (green tones) structures.

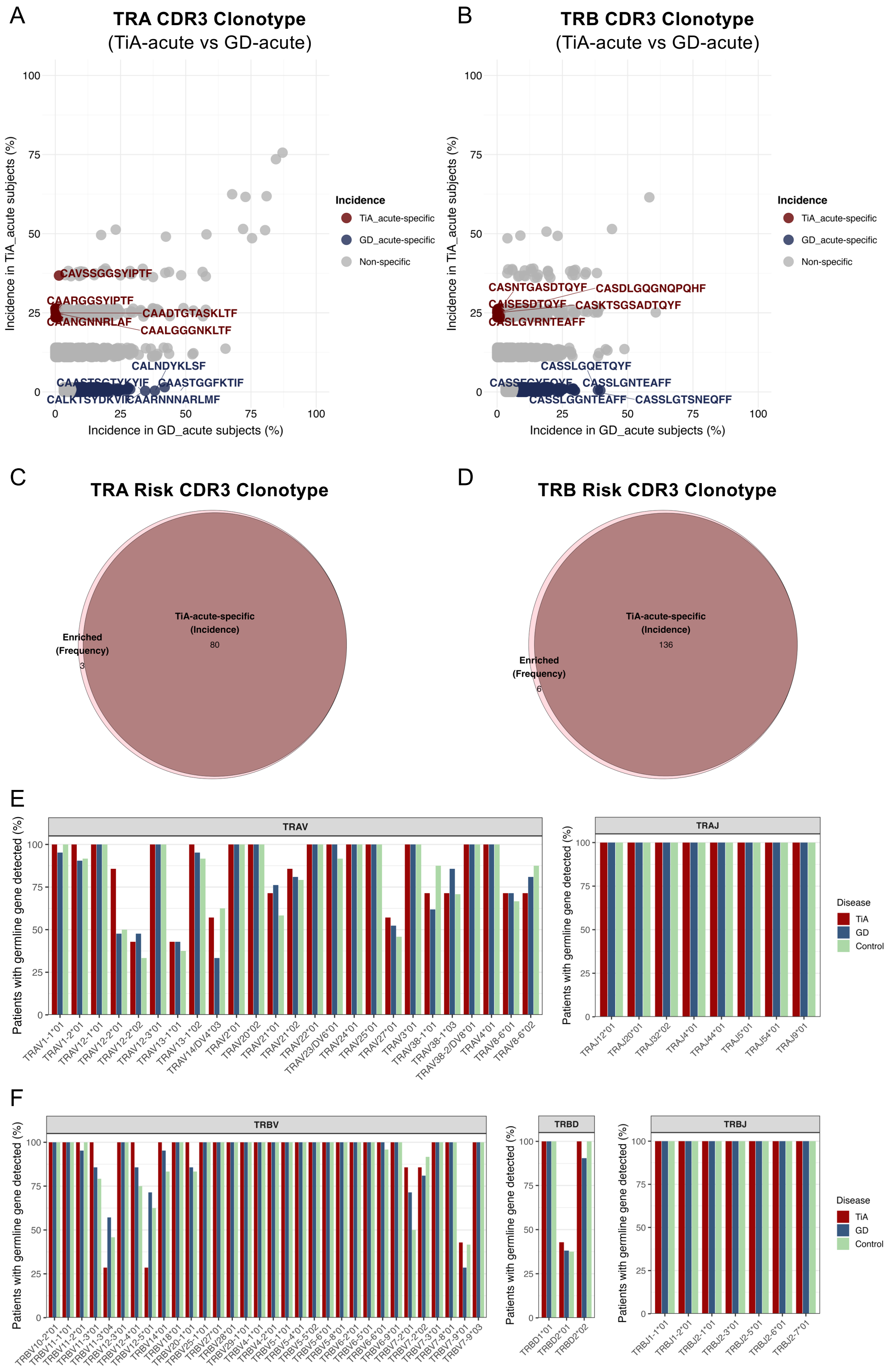


**Figure S5. Further analyses for TiA-associated TCR risk clonotypes.**

**(A-B)** Bubble plots illustrating the clonotype incidence in TiA-acute patients versus GD-acute patients. Dark red dots indicate clonotypes present exclusively in TiA-acute patients and absent in all GD-acute subjects.; dark blue dots indicate clonotypes present exclusively in GD-acute subjects and absent in all TiA-acute patients. Gray dots represent clonotypes without significant incidence differences. (**C-D**) Venn diagrams showing the overlap between frequency-based and incidence-based TiA-associated risk clonotypes (C; enriched in TiA-acute or exclusive to TiA-acute patients) and protective clonotypes (D; decreased in TiA-acute or exclusive to GD-acute subjects). Numbers indicate clonotype counts in each category. (**E-F**) Bar plots showing the percentage of patients with germline gene detection for V/D/J genes composing TiA-associated TRA (E; TRAV: left, TRAJ: right) and TRB (F; TRBV: left, TRBD: middle, TRBJ: right) risk clonotypes, across TiA, GD, and control subjects.

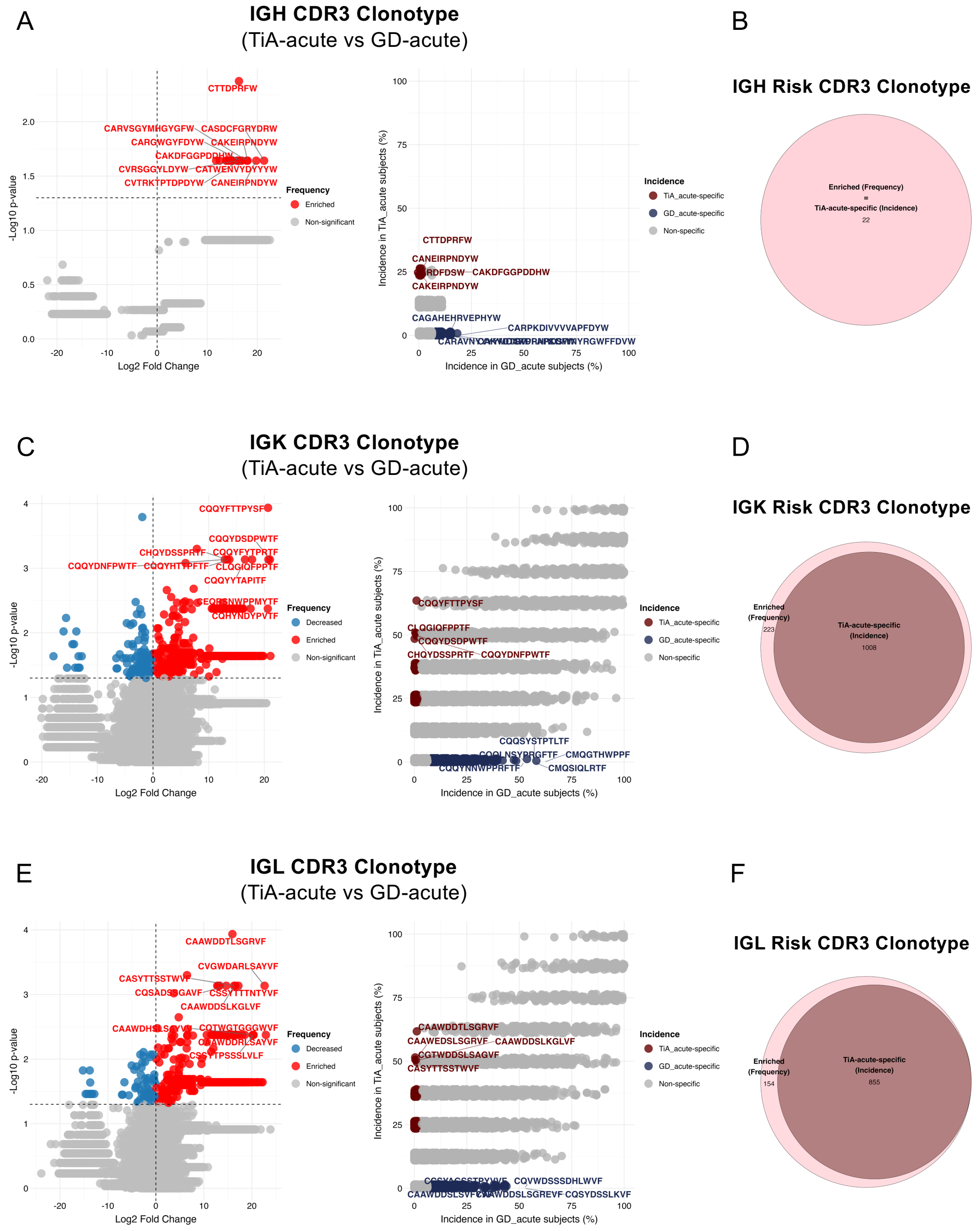


**Figure S6. Identification of TiA-associated BCR risk clonotypes.**

(**A**) Volcano plot (left) and incidence scatter plot (right) of IGH CDR3 clonotypes in TiA-acute versus GD-acute patients. In the volcano plot, red dots: enriched clonotypes ($p < 0.05$, $Log_2FC > 0$); blue dots: decreased clonotypes ($p < 0.05$, $Log_2FC < 0$); gray dots: non-significant clonotypes (Wilcoxon rank-sum test). In the incidence plot, dark red dots: TiA-acute-specific clonotypes (absent in all GD-acute patients); dark blue dots: GD-acute-specific clonotypes (absent in all TiA-acute patients); gray dots: non-specific clonotypes. CDR3 sequences of representative clonotypes are labeled. (**B**) Venn diagram showing the overlap between frequency-based and incidence-based IGH risk clonotypes. (**C-D**) Same as (A-B) for IGK clonotypes. (**E-F**) Same as (A-B) for IGL clonotypes.
